# Metabolome-wide Mendelian randomization characterizes heterogeneous and shared causal effects of metabolites on human health

**DOI:** 10.1101/2023.06.26.23291721

**Authors:** Xianyong Yin, Jack Li, Debraj Bose, Jeffrey Okamoto, Annie Kwon, Anne U. Jackson, Lilian Fernandes Silva, Anniina Oravilahti, Heather M. Stringham, Samuli Ripatti, Mark Daly, Aarno Palotie, Laura J. Scott, Charles F. Burant, Eric B. Fauman, Xiaoquan Wen, Michael Boehnke, Markku Laakso, Jean Morrison

## Abstract

Metabolites are small molecules that are useful for estimating disease risk and elucidating disease biology. Nevertheless, their causal effects on human diseases have not been evaluated comprehensively. We performed two-sample Mendelian randomization to systematically infer the causal effects of 1,099 plasma metabolites measured in 6,136 Finnish men from the METSIM study on risk of 2,099 binary disease endpoints measured in 309,154 Finnish individuals from FinnGen. We identified evidence for 282 causal effects of 70 metabolites on 183 disease endpoints (FDR<1%). We found 25 metabolites with potential causal effects across multiple disease domains, including ascorbic acid 2-sulfate affecting 26 disease endpoints in 12 disease domains. Our study suggests that N-acetyl-2-aminooctanoate and glycocholenate sulfate affect risk of atrial fibrillation through two distinct metabolic pathways and that N-methylpipecolate may mediate the causal effect of N6, N6-dimethyllysine on anxious personality disorder. This study highlights the broad causal impact of plasma metabolites and widespread metabolic connections across diseases.

## Introduction

Metabolites are intermediate or end products of cellular metabolism with a wide range of functions^1^. Compared to gene transcripts and proteins, metabolites are more proximal to diseases, making them ideal biomarkers for estimating disease risk and understanding disease biology. Metabolite levels have shown associations with many human diseases, including type 2 diabetes and multiple cancers^2, 3^. Some metabolites have demonstrated potential for predicting future disease^4, 5^. However, the causal effects of metabolites on human diseases have not been evaluated comprehensively.

Metabolite levels reflect both environmental and genetic influences^1^. With the advent of high-throughput metabolic profiling technology, measuring levels of thousands of metabolites for participants in population studies has become possible. Recent genome-wide association studies (GWAS) that combine high-throughput metabolic profiling and genotyping/sequencing in large samples have identified thousands of genetic associations for thousands of metabolites and metabolic features^6^. These studies usually measure metabolite levels in blood, which are widely considered to reflect metabolite aggregate concentrations across tissues^7^. Recently, we profiled plasma levels for 1,391 metabolites using Metabolon non-targeted mass spectrometry technology in 6,136 Finnish individuals of the Metabolic Syndrome in Men (METSIM) study^8^. GWAS identified 2,030 genetic associations for 803 of the 1,391 metabolites^8^. Integrating these metabolite GWAS with expression quantitative trait loci (eQTL) in 49 human tissues established associations of expression levels of 397 genes with levels of 521 plasma metabolites^9^. These GWAS deepen our understanding of genetic regulation of metabolic individuality, open an avenue to evaluate the causal effects of blood metabolites on human diseases using Mendelian randomization, and have the potential to provide actionable disease interventions.

Mendelian randomization is an instrumental variable (IV) method to interrogate causal effects of heritable risk factors on diseases of interest using genetic variants as IVs^10^. Mendelian randomization has identified modifiable risk factors for human diseases and recent methods development facilitates its broader application. For example, Mendelian randomization using the robust adjusted profile score (MR-RAPS) can account for bias of weak and outlier genetic IVs^11^ and multivariable Mendelian randomization enables testing causal effects of multiple potentially related exposures on the same outcome^12, 13^.

Mendelian randomization analysis has recently been applied to search for causal blood metabolites for a wide range of diseases and traits, including type 2 diabetes^14^, neuroticism^15^, Alzheimer’s disease^16^, and rheumatoid arthritis^17^. These studies demonstrate the utility of Mendelian randomization to identify potential causal metabolites and metabolic pathways for human diseases. However, the existing studies are restricted to one or a few disease outcomes and a relatively limited set of metabolites^6, 18^.

Here, we comprehensively evaluated potential causal effects of 1,099 plasma metabolites on 2,099 binary disease endpoints (hereafter disease traits) using a Mendelian randomization analysis in GWAS of METSIM plasma metabolites^8^ and FinnGen disease traits (release 7)^19^. We identified evidence for 282 causal effects of 70 plasma metabolites on 183 disease traits. Our study uncovered new potential causal effects of plasma metabolites for a broad spectrum of human diseases. We also identified some metabolites with broad causal effects across multiple disease types.

## Results

### Interpretation of Mendelian randomization effect estimates

Mendelian randomization tests whether genetic variants that affect the exposure (metabolite) have a proportional effect on the outcome (disease trait). With additional assumptions about the relationship between the genetic variants, metabolites, and disease traits^20^, the proportionality constant can be interpreted as a measure of the strength of the causal effect. In this paper, we focus primarily on significance and direction when interpreting estimated effects. Mendelian randomization can avoid bias due to environmental confounding and reverse causation which can plague observational associations^20^. However, causal interpretation of Mendelian randomization effects relies on additional assumptions, which may not hold in all cases. These effects must therefore be interpreted in the context of other sources of evidence (see Davies et al. 2017^20^ for a full discussion of interpretation of Mendelian randomization estimates).

### Summary of Mendelian randomization results

We previously conducted GWAS for 1,099 named plasma metabolites with annotated chemical identities in up to 6,136 Finnish men aged 45-74 at enrollment from the METSIM study^8^. These 1,099 metabolites included nine biochemical classes of small molecules related to the metabolisms of lipids (n=548, 49.9%), amino acids (n=215, 19.6%), xenobiotics (n=163, 14.8%), peptides (n=42, 3.8%), nucleotides (n=42, 3.8%), cofactors and vitamins (n=38, 3.5%), carbohydrates (n=25, 2.3%), partially-characterized molecules (n=16, 1.5%), and energy (n=10, 0.9%) (**Supplementary Table 1**).

To identify causal plasma metabolites for human diseases, we carried out univariable Mendelian randomization analysis using MR-RAPS^19^ to evaluate causal effects of the 1,099 metabolites on 2,099 binary disease traits from the FinnGen study (release 7; **Fig. 1a**). In GWAS, we inverse normalized the metabolite measurements^8^ and measured disease trait associations by mixed-model logistic regression^19^. Our estimated causal effects can therefore be interpreted as the change in log odds of disease risk caused by an increase of one standard deviation of the normalized metabolite level. To identify independent IVs for the Mendelian randomization analysis, we performed linkage disequilibrium (LD) clumping in the GWAS summary statistics for each of the 1,099 metabolites to ensure resulting variants achieve association *P* < 10^-5^ and each pair of variants within 1 megabase (Mb) distance satisfy LD r^2^ < 0.01. For the 1,099 metabolites, we identified from 12 to 173 likely independent variants (mean=42.3; median=40.0) and used these as IVs (**Supplementary Fig. 1**).

**Figure 1:**
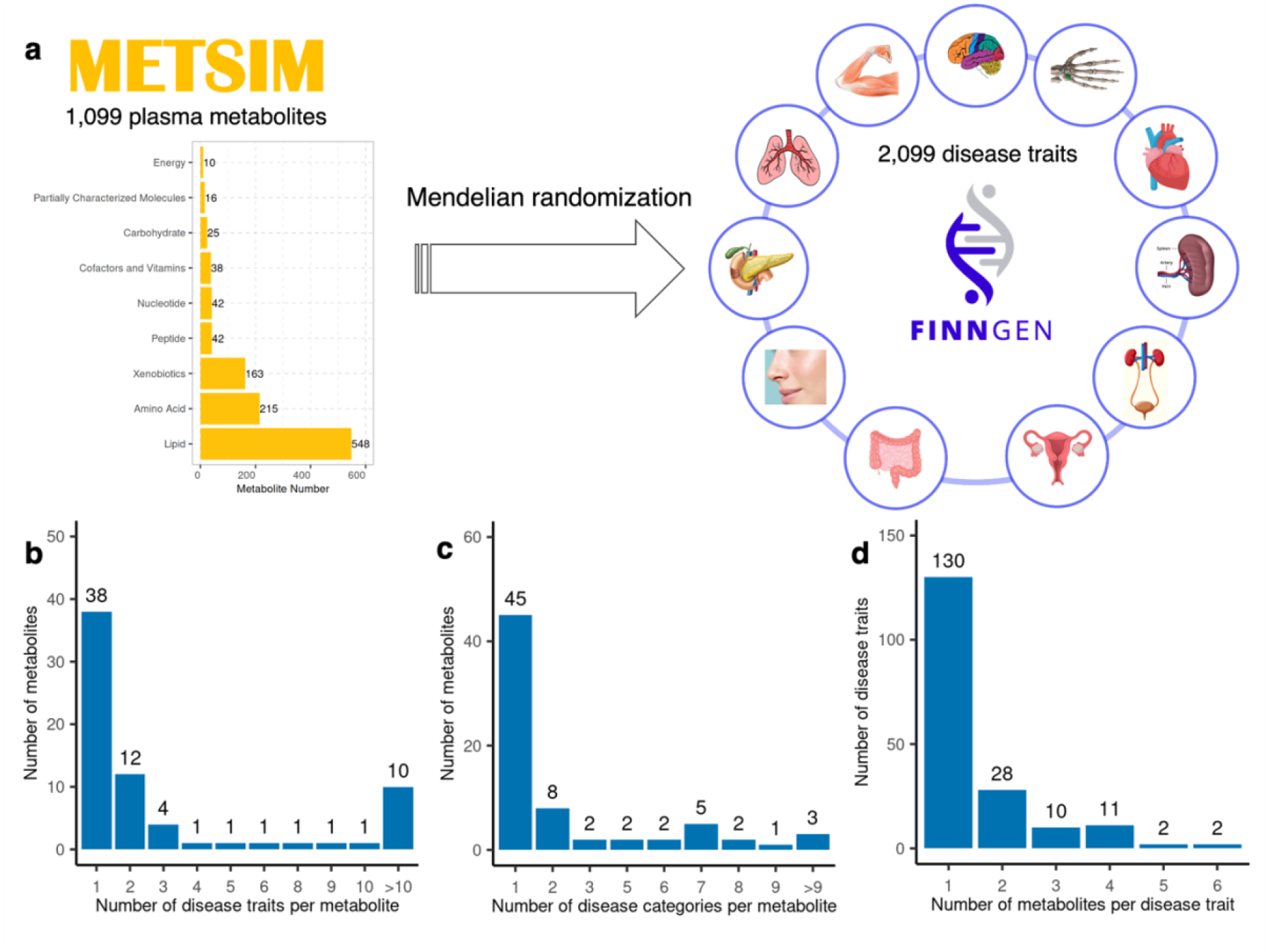
Summary of the 282 significant causal effects of 70 metabolites on 183 disease traits. **a**, the overall design of univariable Mendelian randomization to test causal effects of 1,099 metabolites on 2,099 disease traits; **b**, distribution of metabolites by the number of disease traits that they showed significant causal effects on; **c**, distribution of metabolites by the number of disease categories that they showed significant causal effects on; **d**, distribution of disease traits by the number of their associated causal metabolites.

We identified evidence for 282 causal effects of 70 plasma metabolites on 183 disease traits at a false discovery rate (FDR) threshold < 1% (**Fig. 2** and **Supplementary Table 2**), highlighting the broad relevance of plasma metabolite levels to human health. These 282 metabolite-disease trait pairs showed strong robustness to IV selection and choice of Mendelian randomization method (**Supplementary Fig. 2-5, Supplementary Notes**). The 70 causal metabolites comprised lipids (n=31, 44.3%), amino acids (n=29, 41.4%), xenobiotics (n=4, 5.7%), cofactors and vitamins (n=2, 2.9%), and nucleotides, carbohydrate, peptide, and partially-characterized molecule (n=1, 1.4% for each). Compared to the 1,099 metabolites evaluated, the 70 metabolites showed significant enrichment in amino acids (odds ratio (OR)=3.20, Chi-square test *P*=4.0×10^-6^) and depletion in xenobiotics (OR=0.33, Chi-square test *P*=0.041), which may reflect the significantly larger numbers of IVs for amino acids than for xenobiotics (Student’s t-test *P*=1.2×10^-12^). The 70 plasma metabolites conferred significant causal effects on 1 to 26 disease traits (mean=4.0; median=1.0), with 32 (46%) showing significant causal effects on more than one disease trait (**Fig. 1b-1c**). The 183 disease traits covered a broad spectrum of diseases. The FinnGen consortium grouped these disease traits into 20 categories, including cancers (e.g. colon cancers), cardiometabolic (e.g. type 2 diabetes), infectious (e.g. tularaemia), neurological (e.g. Parkinson’s disease), and mental and behavioral diseases (e.g. anxiety personality disorder) (**Supplementary Table 2**). Each of the 183 disease traits had 1 to 6 causal metabolites (mean=1.5; median=1.0); 53 (29%) had ≥2 causal metabolites (**Fig. 1d**).

**Figure 2:**
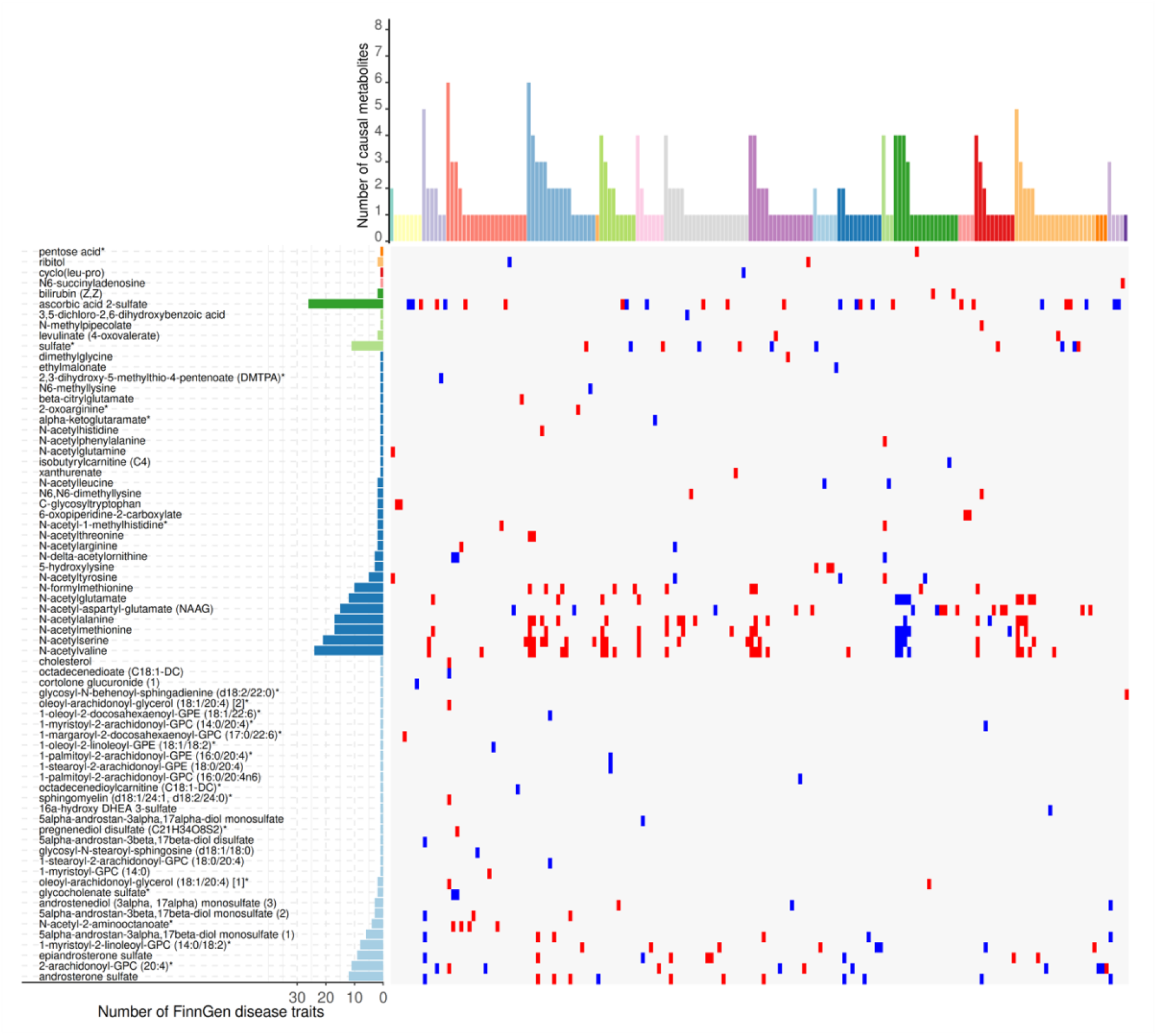
Heat map of the 282 potential causal effects of 70 metabolites on 183 FinnGen disease traits. The x-axis denotes the 183 disease traits of 20 colored categories (from left to right): light paleturquoise (n=1; alcohol related diseases), light wheat (7; congenital malformations, deformations and chromosomal abnormalities), light steel blue (6; diseases of the blood and blood-forming organs), salmon (20; diseases of the circulatory system), sky blue (17; diseases of the digestive system), dark sandy brown (1; diseases of the ear and mastoid process), dark olive green (9; diseases of the eye and adnexa), light thistle (7; diseases of the genitourinary system), gray (21; diseases of the musculoskeletal system and connective tissue), orchid (16; diseases of the nervous system), light sky blue (6; diseases of the respiratory system), dodger blue (11; diseases of the skin and subcutaneous tissue), dark sea green (3; drug purchase endpoints), forest green (16; endocrine, nutritional and metabolic diseases), light pink (4; infectious and parasitic diseases), fire brick (10; mental and behavioral disorders), sandy brown (20; neoplasms), dark orange (3; neurological diseases), medium thistle (4; pregnancy, childbirth and the puerperium), and medium purple (1; rheuma endpoints). The y-axis denotes the 70 metabolites of eight colored biochemical classes (from bottom to top): light blue (n=31; lipids), dark blue (29; amino acids), light green (4; xenobiotics), dark green (2; cofactors and vitamins), pink (1; nucleotides), red (1; peptides), light orange (1; carbohydrates), and dark orange (1; partially characterized molecules). The bar plots show the number of FinnGen disease traits that each metabolite confers causal effects on (on the left) and the number of causal metabolites for each disease trait (on the top). The color of cells denotes the direction of potential causal effects (red for positive and blue for negative effects) of metabolites on disease traits.

### New potential causal metabolites for diseases

Among the 282 causal effects, we reproduced several known relationships. For example, we identified a potential causal effect of low plasma lipid glycosyl-N-stearoyl-sphingosine levels on increasing risk of coronary artery disease (β=-0.11, *P*=1.0×10^-6^), reinforcing the important role of sphingolipid metabolism in coronary artery disease^21^. Studies have reported high levels of valine, a branched-chain amino acid, associated with increased risk of type 2 diabetes^4, 22^. We validated, with nominal significance, the causal effect of plasma valine levels on risk of type 2 diabetes (β=0.041, *P*=5.0×10^-3^). In addition, we found that elevated plasma N-acetylvaline levels decreased risk of type 2 diabetes (β=-0.085, *P*=1.1×10^-8^). N-acetylvaline is a derivative of valine and belongs to a class of N-acyl-alpha amino acids. Multivariable Mendelian randomization including both valine and N-acetylvaline suggested that both metabolites have direct effects on type 2 diabetes (N-acetylvaline: β=-0.096, *P*=2.7×10^-12^; valine: β=0.087, *P*=1.8×10^-5^), indicating a potentially important and complex role of valine metabolism in risk of type 2 diabetes. Interestingly, we found that high levels of two additional plasma N-acyl-alpha amino acids N-acetylglutamate (β=-0.11, *P*=1.0×10^-7^) and N-acetylmethionine (β=-0.072, *P*=5.5×10^-7^) potentially causally decreased risk of type 2 diabetes. The three N-acyl-alpha amino acids N-acetylvaline, N-acetylglutamate, and N-acetylmethionine show substantial phenotypic correlation and share many IVs (**Fig. 3**, **Supplementary Fig. 6**). For these three N-acyl-alpha amino acids, our GWAS previously identified genome-wide significant associations at the *ACY1* gene^8^, which encodes enzyme aminoacylase 1 that catalyzes the hydrolysis of acylated L-amino acids to L-amino acids. Mendelian randomization suggested that elevated plasma aminoacylase 1 levels^23^ decreased levels of the three N-acyl-alpha amino acids (β<-1.20, *P*<4.2×10^-21^) but increased risk of type 2 diabetes (β=0.16, *P*=2.6×10^-4^), directionally consistent with the known function of aminoacylase 1 and a recently reported positive causal effect of aminoacylase 1 on type 2 diabetes^24^. These findings suggest a possible role of synthesis or degradation of N-acetylated proteins in type 2 diabetes. However, due to substantial sharing of IVs across the three N-acetyl amino acids, Mendelian randomization cannot identify whether this effect is due to one specific N-acetyl amino acid or multiple.

**Figure 3:**
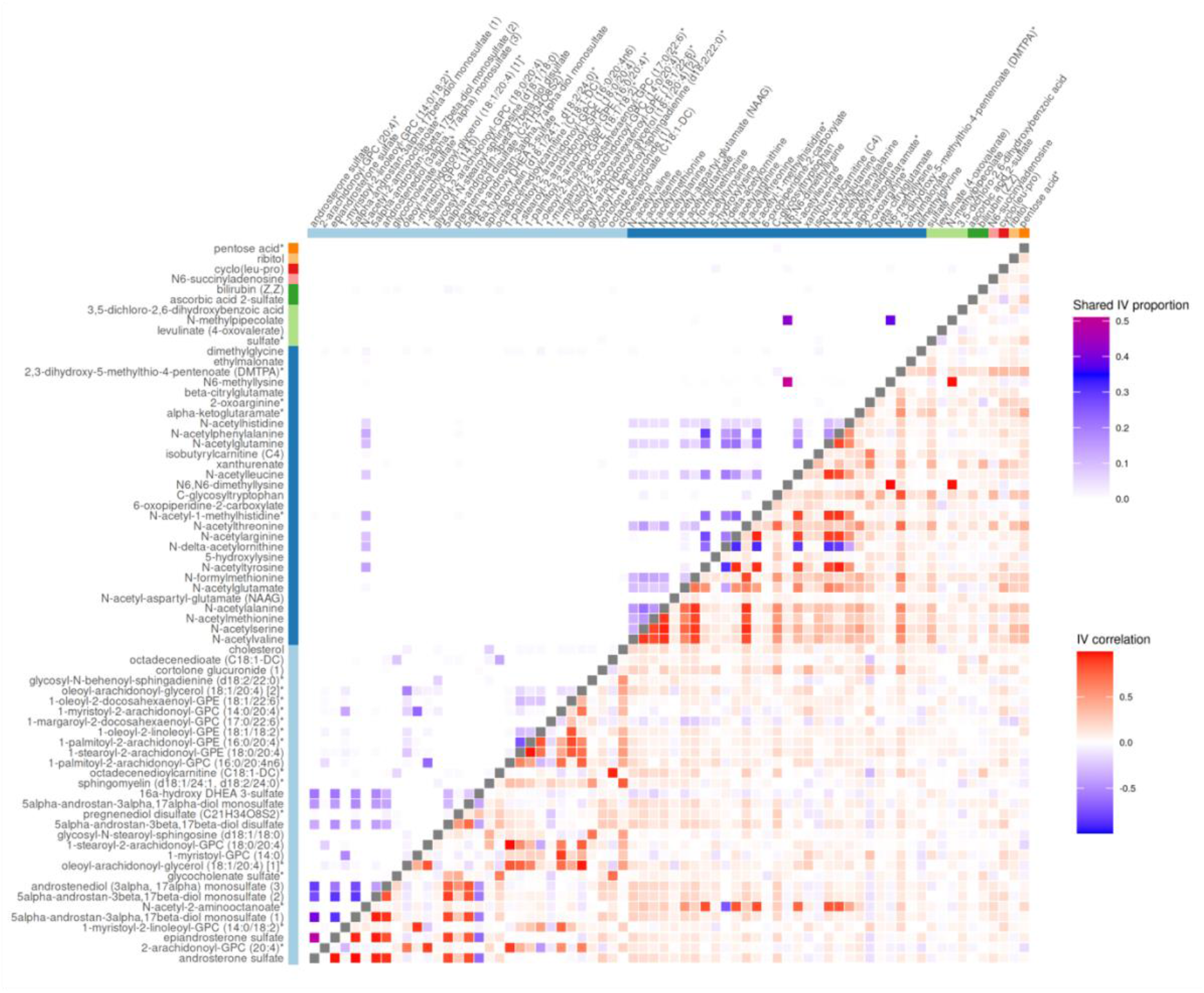
IV sharing (upper left triangular heat map) and correlation (lower right triangular heat map) for all pairs of the 70 metabolites. The color bar on the x-axis and y-axis denotes the biochemical classes of metabolites: light blue (lipids), dark blue (amino acids), light green (xenobiotics), dark green (cofactors and vitamins), pink (nucleotides), red (peptides), light orange (carbohydrates), and dark orange (partially characterized molecules). In the upper left triangular heat map, each cell denotes the proportion of IVs with metabolite association *P*≤10^-5^ shared between the pair of metabolites. In the lower right triangular heat map, and each cell denotes the IV correlation between the pair of metabolites. The diagonal cells are colored in dark gray to distinguish the upper and lower triangular heat maps.

Our study also identified new potential causal metabolites for human diseases. Mendelian randomization recently suggested causal effects of plasma metabolites on risk of dementia^16, 25, 26^. We identified a significant potential protective effect of high plasma lipid 2-arachidonoyl-GPC (20:4) levels on risk of frontotemporal dementia (β=-0.89, *P*=1.2×10^-6^), a type of dementia characterized by progressive loss of neurons in the brain’s frontal or temporal lobes. To the best of our knowledge, studies previously only reported 2-methoxyacetaminophen sulfate^27^ with causal effect specifically on frontotemporal dementia. 2-arachidonoyl-GPC (20:4) is a lysophosphatidylcholine widely considered as a potent pro-inflammatory mediator^28^. Emerging evidence has demonstrated that neuroinflammation plays an important role in dementia^29^. Studies have identified a negative association of lysophosphatidylcholine with Alzheimer’s disease^30^. Consistent with these results, we found a protective causal effect of increased 2-arachidonoyl-GPC (20:4) levels on risk of frontotemporal dementia. We previously identified genome-wide associations for 2-arachidonoyl-GPC (20:4) around the *FADS1/FADS2,* two fatty acid desaturase genes^8^. Interestingly, we found that low expression of *FADS1/FADS2* in the whole blood but high expression in the brain significantly increased plasma 2-arachidonoyl-GPC (20:4) level^9^. *FADS1* variants could regulate erythrocyte arachidonic acid biosynthesis that subsequently induces inflammation in Alzheimer’s disease^31^.

Chronic kidney disease affects >10% of the general population worldwide^32^ and its risk factors are still poorly understood. We found evidence that elevated plasma xenobiotic sulfate levels increased risk of chronic kidney disease (β=0.080, *P*=1.9×10^-7^). High sulfate levels have been previously found to be associated with disease progression and increased mortality in individuals with kidney disease^33^. Our previous GWAS identified a genome-wide significant association with plasma sulfate levels at the *SLC13A1* gene^8^, which encodes a sulfate transmembrane transporter and mediates the first step of sulfate absorption. *SLC13A1* is primarily expressed in the proximal renal tubules. We previously found that high expression of *SLC13A1* decreased plasma sulfate abundance^9^. These results together suggest that *SLC13A1* could serve as a potential drug target for chronic kidney disease through regulation of plasma sulfate levels.

### Causal metabolites shared across diseases

We identified evidence for 32 metabolites with causal effects on more than one disease trait (**Fig. 1b** and **Fig. 2**; see **Summary of Mendelian randomization results**). Of these 32 metabolites, 25 (78%) showed significant causal effects on ≥2 distinct disease categories (**Fig. 1c**, **Supplementary Table 2**). The sharing of causal metabolites between diseases may partially explain observed phenotypic correlations and disease comorbidities. For example, we identified causal effects of plasma amino acid N-acetylvaline levels on optic atrophy (β=0.53, *P*=4.7×10^-7^) and myasthenia gravis (β=0.53, *P*=7.9×10^-8^), diseases with substantial comorbidity^34^. These results suggested that valine metabolism might play a role in both cell cycle of retinal ganglion cell axons and communication between nerves and muscle. We found causal effects of plasma amino acid N-acetyl-aspartyl-glutamate (NAAG) levels on increased risk of both Parkinson’s disease (β=0.11, *P*=3.2×10^-7^) and autoimmune hypothyroidism (β=0.039, *P*=3.9×10^-9^), which also have substantial comorbidity^35^. To the best of our knowledge, this is the first reported evidence of these four causal effects.

The metabolite affecting the largest number of disease traits was ascorbic acid 2-sulfate, with evidence of causal effects on 26 disease traits of 12 categories, including cardiomyopathy (disease of the circulatory system), arthropathy (disease of the musculoskeletal system and connective tissue), and acne (disease of the skin and subcutaneous tissue) (**Supplementary Table 2**). We found that elevated levels of ascorbic acid 2-sulfate may decrease risk of 12 disease traits including colon adenocarcinoma (β=-0.13; *P*=9.3×10^-8^) and endometriosis of the fallopian tube (β=-0.48; *P*=1.6×10^-7^) but increase risk of 14 others including conjunctiva cancer (β=0.36; *P*=2.8×10^-14^) and arthropathy (β=0.028; *P*=1.1×10^-7^).

Notably, the suggested causal effects of plasma ascorbic acid 2-sulfate showed heterogeneity across disease traits even in the same category. For example, we found elevated ascorbic acid 2-sulfate levels are protective for acne (β=-0.18; *P*=3.9×10^-10^) and lichen sclerosus (β=-0.15; *P*=7.1×10^-7^) but increase risk of dyshidrosis, a kind of eczema (β=0.42; *P*=4.2×10^-10^). These three conditions all affect skin but usually in different anatomical locations: the face, upper part of the chest, and back; the genital area; and the palms and fingers, respectively. Ascorbic acid 2-sulfate arises from the action of a liver-derived sulfotransferase on vitamin C, so it is possible that plasma levels of ascorbic acid 2-sulfate are a proxy for action of liver-derived sulfotransferases or for vitamin C levels, or a combination of these. Vitamin C is an essential nutrient for humans, acting as an antioxidant by protecting the body against oxidative stress, as a cofactor in enzymatic reactions including collagen synthesis, and as a structure component for blood vessels, cartilage, and muscle^36^. Vitamin C supplementation has been broadly recommended to help protect cells against the effects of free radicals, and has generally been found to be safe. Further investigation is needed to understand whether the effects we identified are effects of vitamin C itself or other biological processes.

### Independent causal metabolic pathways on the same disease

We computed both phenotypic correlation and correlation of IV effects (r_IV_) for each pair of the 70 significant metabolites, showing their pervasive connections (**Fig. 3**, **Supplementary Fig. 7-8**; see **Methods**). We found strong correlations between some pairs of potential causal metabolites for the same disease traits (absolute r_IV_ median=0.84, mean=0.64, range=0.00033-0.99; **Supplementary Fig. 9**).

Causal effects of two metabolites with highly correlated IVs on the same disease trait in univariable Mendelian randomization could result from multiple scenarios. For example, both metabolites may causally affect the disease trait independently, or only one metabolite could affect the disease trait, with the result for the other being due to mediation or horizontal pleiotropy. We employed multivariable Mendelian randomization^13^ to distinguish these possibilities.

For atrial fibrillation, we identified a risk effect of plasma lipid N-acetyl-2-amino-octanoate (β=0.068; *P*=2.3×10^-7^) and protective effects of plasma amino acid N-delta-acetylornithine (β=-0.047; *P*=5.1×10^-7^) and lipid glycocholenate sulfate (β=-0.061; *P*=2.9×10^-8^). N-acetyl-2-aminooctanoate and N-delta-acetylornithine have highly correlated IVs (r_IV_=0.74) but neither has correlated IVs with glycocholenate sulfate (|r_IV_|<0.08). Multivariable Mendelian randomization analysis identified distinct causal effects on atrial fibrillation of lipids N-acetyl-2-amino-octanoate (β=0.054; *P*=7.2×10^-3^) and glycocholenate sulfate (β=-0.058; *P*=2.6×10^-7^), but no causal effect of N-delta-acetylornithine, conditional on the other two metabolites (β=-0.020; *P*=0.17). In the METSIM study, we identified 816 individuals with atrial fibrillation (see **Methods**). Logistic regression identified a significant association between plasma N-acetyl-2-amino-octanoate level and risk of atrial fibrillation (β=0.080; *P*=0.045), directionally consistent with the causal effect estimated in Mendelian randomization. We observed no significant associations with N-delta-acetylornithine (β=0.057; *P*=0.148) or glycocholenate sulfate levels (β=0.072; *P*=0.064), however observational associations may be biased by unmeasured confounding variables.

For anxious personality disorder, we identified risk effects of plasma xenobiotic N-methylpipecolate (β=0.28; *P*=2.8×10^-7^) and amino acid N6, N6-dimethyllysine (β=0.24; *P*=8.6×10^-8^) and a protective effect of plasma lipid androsterone sulfate (β=-0.27; *P*=1.5×10^-7^). N6,N6-dimethyllysine and N-methylpipecolate have high IV correlation (r_IV_=0.98) and share 42.4% of their IVs at a threshold of metabolite association *P*≤1×10^-5^, but neither has correlated IVs with androsterone sulfate (|r_IV_|<0.03). Because of the high IV correlation between N6, N6-dimethyllysine and N-methylpipecolate, there is not enough independent genetic signal to tease apart their causal effects on anxious personality disorder using multivariable Mendelian randomization. We performed two multivariable Mendelian randomization analyses including androsterone sulfate and either N-methylpipecolate or N6, N6-dimethyllysine. In both cases, the data to be consistent with direct effects of both included metabolites (N-methylpipecolate (β=0.29; *P*=6.2×10^-8^) and androsterone sulfate (β=-0.27; *P*=7.6×10^-8^) or at N6, N6-dimethyllysine (β=0.24; *P*=5.0×10^-7^) and androsterone sulfate (β=-0.27; *P*=2.5×10^-7^)). To further understand this relationship, we carried out a GWAS on the metabolite ratio of N6,N6-dimethyllysine and N-methylpipecolate, identifying six independent association signals in the *AKR1C1/AKR1C2/AKR1C3/AKR1C4/AKR1C8, NAT8, PYROXD2, SLC6A20*, and *SLC7A9* regions (*P*<5.0×10^-8^) (**Supplementary Table 3**, **Supplementary Fig. 10**). Mendelian randomization identified evidence for a causal effect of increased N6,N6-dimethyllysine:N-methylpipecolate ratio on risk of anxious personality disorder (β=-0.34; *P*=0.047; **Supplementary Fig. 11**; see **Methods**). The pattern we observe in which N6, N6-dimethyllysine and N-methylpipecolate both increase risk of anxious personality disorder, but an increase in their ratio confers a protective effect is consistent with a hypothesis that N-methylpipecolate acts as a mediator in the potential causal pathway of N6, N6-dimethyllysine on anxious personality disorder (**Fig. 4**). This is consistent with previous reports that pipecolate is an intermediate product of lysine metabolism^37^.

**Figure 4:**
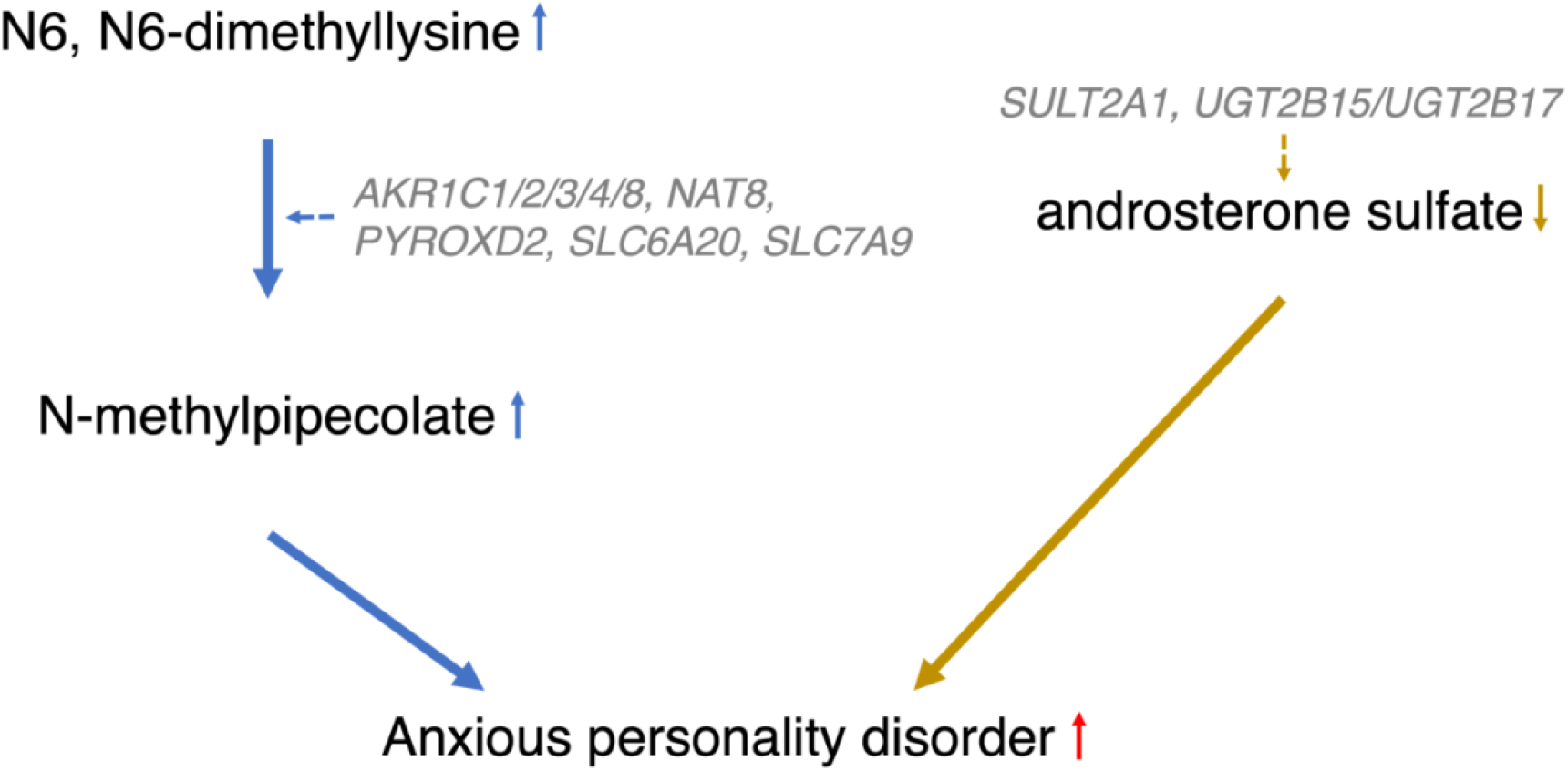
Mendelian randomization suggests two metabolic pathways for anxious personality disorder. Genes implicated for the ratio of N6,N6-dimethyllysine and N-methylpipecolate and for androsterone sulfate are italicized.

## Discussion

In this study, we systematically screened for potential causal effects of 1,099 plasma metabolites on 2,099 disease endpoints using two-sample univariable and multivariable Mendelian randomization analysis. We identified evidence for 282 causal effects of 70 plasma metabolites on 183 disease endpoints. We characterized the sharing of metabolite causal effects across 53 human diseases and showed the heterogeneity of causal metabolic pathways in disease pathophysiology. This study uncovers modifiable risk metabolites for disease intervention and underscores a pervasive potential causal role of plasma metabolites in human health.

We identified evidence for causal effects of 70 plasma metabolites on 183 human diseases. The relationships of many plasma metabolites with diseases have not been studied previously. These findings have several implications. First, they provide potential targets for disease intervention. Many plasma metabolites levels can be modified by diet and lifestyle changes. For example, we identified that high plasma sulfate levels increased risk of chronic kidney disease. A wide range of food and beverages have been suggested as sources of dietary sulfate. We can, in principle, reduce plasma sulfate levels by reducing the consumption of these food and beverages.

Second, these findings help elucidate disease biology and prioritize therapeutic targets for human diseases. For example, the risk of high plasma sulfate on chronic kidney disease suggested *SLC13A1* as a potential drug target for chronic kidney disease. The protective effect of high 2-arachidonoyl-GPC (20:4) level on frontotemporal dementia bolsters the hypothesis that neuroinflammation contributes to the pathophysiology of dementia^29, 31^. We characterize the pervasive sharing of potential causal metabolites and their heterogeneity effects across human diseases. The sharing may help explain some disease comorbidity and reveal previously-unappreciated connections between diseases. For example, we identified evidence for 126 heterogeneous causal effects of 15 N-acyl-alpha amino acids on 67 disease traits of 14 categories, highlighting a broad impact of synthesis or degradation of N-acetylated proteins on human health.

Our study showed that metabolites with significant univariable causal effects on the same disease traits might act in disease pathogenesis through separate metabolic pathways or through a metabolic cascade. We identified two independent metabolic pathways among three tested metabolites for atrial fibrillation and for anxious personality disorder, highlighting the heterogeneity of potential causal metabolic pathways in human diseases. We suggested that a causal effect of N-delta-acetylornithine on atrial fibrillation might be induced by IVs shared with N-acetyl-2-amino-octanoate. In contrast, we suggested that N-methylpipecolate might act as a downstream mediator in the causal pathway of N6, N6-dimethyllysine on anxious personality disorder, which could partially explain the strong IV correlation between N-methylpipecolate and N6, N6-dimethyllysine. Previous survival analyses detected significant positive association of glycocholenate sulfate levels with atrial fibrillation incidence^38^, while our analysis identified a negative association of plasma glycocholenate sulfate with atrial fibrillation. The effect of plasma glycocholenate sulfate on atrial fibrillation warrants further investigation.

In this study, plasma metabolite levels may act as proxies for activity of specific biological pathways, or levels of metabolites in other tissues. For ease of exposition, we referred to causal effects of metabolites on disease traits throughout this report. However, this may not mean that intervening directly on plasma metabolite levels will impact risk of disease trait. The biochemical pathway regulating the metabolite may be the true causal culprit. For example, we identified a negative association of plasma 2-arachidonoyl-GPC (20:4) level with the risk of frontotemporal dementia, which might suggest a role of 2-arachidonoyl-GPC (20:4)-mediated neuroinflammation in the brain. In addition, we applied multivariable Mendelian randomization to tease apart independent potential causal effects of metabolites on the same disease. However, multivariable Mendelian randomization can only distinguish effects of metabolites that have a sufficient number of distinct IVs. Finally, associations between IVs and metabolites were estimated in an all-male cohort^8^, while IV-disease associations were estimated in a mixed cohort. Our causal effect estimates rely on the assumption that genetic regulation of metabolites and causal effects do not differ between the sexes. If these assumptions are violated, our estimates will be inaccurate or may not generalize to a mixed sex population. This issue is most likely to affect sexually differentiated metabolites such as androsterone sulfate.

In conclusion, we systematically evaluated the causal effects of 1,099 plasma metabolites on the risk of 2,099 disease endpoints. We identified evidence for 282 causal effects of 70 plasma metabolites on 183 disease traits. Our study newly uncovered potential causal effects of plasma metabolites on a broad spectrum of human diseases. These findings highlight heterogeneous and shared causal effects of plasma metabolites on human diseases.

## Methods

### Metabolic Syndrome In Men (METSIM) metabolomics study

METSIM is a single-site cohort study designed to investigate risk factors for type 2 diabetes and cardiovascular diseases^39^. It includes 10,197 Finnish men from Kuopio aged 45 to 74 years at baseline. We performed non-targeted metabolomics profiling in 6,136 randomly-selected non-diabetic participants using the Metabolon DiscoveryHD4 mass spectrometry platform (Durham, North Carolina, USA) on EDTA-plasma samples obtained after ≥10-hour overnight fast during baseline visits from 2005 to 2010^8^. We completed single-variant GWAS for 1,391 metabolites, which identified 2,030 independent metabolite associations^8^. For this study, we used GWAS summary statistics at 16.2M genotyped or imputed genetic variants for the 1,099 named metabolites with annotated biochemical identities^8^. All METSIM participants provided written informed consent. The Ethics Committee at the University of Eastern Finland and the Institutional Review Board at the University of Michigan approved the METSIM metabolomics study.

### FinnGen study

FinnGen is designed to collect and analyze genome and healthcare data to identify new diagnostic and therapeutic targets for human diseases^19^. FinnGen obtained participant informed consent for biobank research based on the Finnish Biobank Act. Research cohorts collected prior to the Finnish Biobank Act coming into effect (September 2013) and the start of FinnGen (August 2017) obtained study-specific consents and later transferred the consents to the Finnish biobank after the National Supervisory Authority for Welfare and Health (Fimea) approved the recruitment protocols.

FinnGen identified 3,095 disease endpoints in release 7 using healthcare data from Finnish national registries: Drug Purchase and Drug Reimbursement and Digital and Population Data Services Agency; Digital and Population Data Services Agency; Statistics Finland; Register of Primary Health Care Visits (AVOHILMO); Care Register for Health Care (HILMO); and Finnish Cancer Registry. These registries recorded disease-relevant codes of the International Classification of Diseases (ICD) revisions 8, 9, and 10, cancer-specific ICD-O-3, Nordic Medico-Statistical Committee (NOMESCO) procedure, Finnish-specific Social Insurance Institute (KELA) drug reimbursement, and Anatomical Therapeutic Chemical (ATC)^8^. Each FinnGen participant was genotyped with an Illumina or Affymetrix array. Genotype imputation followed using the Finnish-specific Sequencing Initiative Suomi (SISu) v3 reference panel^40^. FinnGen carried out single-variant GWAS for each disease endpoint using mixed model logistic regression in SAIGE^41^. For this study, we used GWAS summary statistics at 16.7M genotyped or imputed genetic variants for all 3,095 disease traits in up to 309,154 individuals from FinnGen release 7. After we finished the Mendelian randomization analysis, FinnGen made the release 8 publicly available, which includes GWAS summary statistics for 2,202 disease traits. In comparison to FinnGen release 7, release 8 reduced the number of disease traits primarily by dropping redundant disease traits. To improve efficiency and reduce redundancy, we restricted our Mendelian randomization analysis results to 2,099 of the 3,095 disease traits that are included in FinnGen release 8.

### Selection of IVs

We identified 16.2M genetic variants shared between GWAS summary files across all the 1,099 metabolites in METSIM and the 2,099 disease traits in FinnGen release 7. To identify independent genetic variants as IVs for Mendelian randomization, we performed LD clumping in the GWAS results for each of the 1,099 metabolites in Plink to ensure resulting variants achieved association *P*<10^-5^ and each pair of variants within 1 Mb distance has LD r^2^<0.01^42^. For LD calculation, we used genotypes in 8,433 METSIM individuals without close relatives defined as pairwise kinship coefficients<0.125.

### Primary univariable Mendelian randomization

To identify causal metabolites for human diseases, we performed two-sample univariable Mendelian randomization to test the causal effect of each of the 1,099 plasma metabolites on each of the 2,099 disease traits using MR–robust adjusted profile scoring (MR-RAPS)^11^. MR-RAPS allows for horizontal pleiotropy and enables inclusion of IVs with weak effects by accounting for the precision of IV-exposure and IV-outcome associations^11^. We used over dispersion and Tukey robust loss function parameters in MR-RAPS. We conducted the MR-RAPS analysis using the mr.raps R package. To identify significant causal effects, we applied an FDR<1% to account for multiple testing.

### Evaluation of causal effects of blood aminoacylase 1 levels on plasma levels of three N-acyl-alpha amino acids and risk of type 2 diabetes

To test causal effects of protein aminoacylase 1 on plasma levels of three N-acyl-alpha amino acids: N-acetylvaline, N-acetylglutamate, and N-acetylmethionine and risk of type 2 diabetes, we performed two-sample univariable Mendelian randomization. deCODE measured plasma aminoacylase 1 level using SomaScan version 4 in 35,559 Icelanders followed by protein quantitative trait loci (pQTL) analysis, which identified three independent cis-pQTLs for aminoacylase 1^23^. Among the three cis-pQTLs, the top pQTL site rs121912698 was available in both METSIM and FinnGen. We used this variant as single IV and performed a Wald ratio test to evaluate causal effects of protein aminoacylase 1 on plasma levels of the three N-acyl-alpha amino acids and risk of type 2 diabetes in the twoSampleMR R package.

### Estimation of IV correlation between metabolites

To estimate the degree to which each pair of metabolites share genetic IVs, we computed the proportion of overlapping IVs and the IV correlation. For each metabolite pair, we took the union of IVs for both metabolites. We then performed LD clumping using LD r^2^<0.01 in 1 Mb distance in Plink^42^ to remove correlated IVs. Finally, we extracted association statistics for the resulting set of IVs for both metabolites. For LD calculation, we used genotypes in 8,433 METSIM individuals with pairwise kinship coefficients<0.125. We calculated the proportion of IVs shared as the proportion of the LD clumped union set of IVs with association *P*≤10^-5^ for both metabolites. We calculated the IV correlation, r_IV_, as the correlation of association statistics of the LD clumped union set of IVs with the two metabolites.

### Multivariable Mendelian randomization

To detect independent causal effects among metabolites that conferred significant univariable causal effects on the same disease trait, we performed multivariable Mendelian randomization in Genome-wide mR Analysis under Pervasive PLEiotropy (GRAPPLE)^13^. We merged the IVs that were used in univariable Mendelian randomization across all the targeted metabolites and performed LD clumping as in **Selection of IVs** to ensure that all IVs were nearly independent. We applied default parameters in GRAPPLE and used nominal *P*<0.05 as the significance threshold.

### Associations of N-acetyl-2-aminooctanoate, N-delta-acetylornithine, and glycocholenate sulfate with atrial fibrillation in METSIM

Among the 6,102 METSIM participants with measured plasma N-acetyl-2-aminooctanoate, N-delta-acetylornithine, and glycocholenate sulfate levels at baseline, we identified 816 with atrial fibrillation in METSIM as of June 2022. To test for associations between plasma metabolite levels and presence of atrial fibrillation, we used logistic regression with covariates baseline study age, body mass index (BMI), binary cigarette smoking status (ever smoker versus never smoker), alcohol drinking amount, baseline systolic and diastolic blood pressure, and lipid and hypertension medication use.

### GWAS for metabolite ratio of N6,N6-dimethyllysine and N-methylpipecolate and causal effect of the ratio on anxious personality disorder

In the 6,136 METSIM participants^8^, we computed the ratio of N6,N6-dimethyllysine to N-methylpipecolate by dividing the level of N6,N6-dimethyllysine by the level of N-methylpipecolate. We regressed out covariates study age, Metabolon batches, and lipid lowering medication status, and inverse normalized the residuals. We performed single-variant GWAS for the resulting residuals in Regenie v3.2.2^43^. For the chromosomes on which we identified genome-wide significant associations (*P*<5.0×10^-8^), we performed recursively a stepwise conditional test to identify near-independent association signals until no variant attained *P*<5.0×10^-8 8^. To test causal effect of the metabolite ratio on risk of anxious personality disorder, we performed univariable Mendelian randomization test using MR-RAPS^11^. We used the near-independent association signals for the metabolite ratio that are also available in the GWAS for anxious personality disorder as IVs. We conducted the MR-RAPS analysis with over dispersion and Tukey robust loss function parameters using the mr.raps R package.

## Data Availability

FinnGen genome-wide summary statistics are available at https://r7.finngen.fi. Full summary statistics from the genome-wide association studies of the 1,099 plasma metabolites are available at https://pheweb.org/metsim-metab/.

https://pheweb.org/metsim-metab/

## Acknowledgements

We thank all the participants and investigators in the METSIM and FinnGen studies. This work was supported by the National Institutes of Health (NIH) under awards U01 DK062370 (M.B.), R35 GM138121 (X.Q.W.), R01 DK119380 (X.Q.W.), the American Diabetes Association Postdoctoral Fellowship (1-19-PDF-061, X.Y.), the University of Michigan Precision Health Scholarship (X.Y.), the Academy of Finland under grant no. 321428 (M.L.), the Sigrid Juselius Foundation (M.L.), the Academy of Finland Center of Excellence in Complex Disease Genetics under grant no. 312062 and 336820 (S.R.), grant no. 312074 and 336824 (A.P.), the Finnish Foundation for Cardiovascular Research (S.R.), University of Helsinki HiLIFE Fellow and Grand Challenge grants, and Horizon 2020 Research and Innovation Programme (grant no. 101016775 “INTERVENE”) (S.R.).

The Coordinating Ethics Committee of the Hospital District of Helsinki and Uusimaa (HUS) approved the FinnGen study protocol Nr HUS/990/2017. The FinnGen study is approved by Finnish Institute for Health and Welfare (permit numbers: THL/2031/6.02.00/2017, THL/1101/5.05.00/2017, THL/341/6.02.00/2018, THL/2222/6.02.00/2018, THL/283/6.02.00/2019, THL/1721/5.05.00/2019, THL/1524/5.05.00/2020, and THL/2364/14.02/2020), Digital and population data service agency (permit numbers: VRK43431/2017-3, VRK/6909/2018-3, VRK/4415/2019-3), Social Insurance Institution (permit numbers: KELA 58/522/2017, KELA 131/522/2018, KELA 70/522/2019, KELA 98/522/2019, KELA 138/522/2019, KELA 2/522/2020, KELA 16/522/2020), and Statistics Finland (permit numbers: TK-53-1041-17 and TK-53-90-20).

## Declaration of interests

E.B.F. is an employee and stockholder of Pfizer. The remaining authors declare no competing interests.

## Web resources

METSIM metabolomics PheWeb: https://pheweb.org/metsim-metab

FinnGen: https://www.finngen.fi

FinnGen documentation: https://finngen.gitbook.io/documentation

## Data and code availability

FinnGen genome-wide summary statistics are available at https://r7.finngen.fi. Full summary statistics from the genome-wide association studies of the 1,099 plasma metabolites are available at https://pheweb.org/metsim-metab/. TwoSampleMR is available at https://github.com/MRCIEU/TwoSampleMR. MR-RAPS is available https://github.com/qingyuanzhao/mr.raps. GRAPPLE is available at https://github.com/jingshuw/GRAPPLE.

## References

1 Wishart, D. S. Metabolomics for Investigating Physiological and Pathophysiological Processes. Physiol Rev 99, 1819–1875 (2019).

2 Ahola-Olli, A. V. et al. Circulating metabolites and the risk of type 2 diabetes: a prospective study of 11,896 young adults from four Finnish cohorts. Diabetologia 62, 2298–2309 (2019).

3 Reznik, E. et al. A Landscape of Metabolic Variation across Tumor Types. Cell Syst 6, 301–313.e303 (2018).

4 Wang, T. J. et al. Metabolite profiles and the risk of developing diabetes. Nat Med 17, 448–453 (2011).

5 Wu, Q. et al. Prediction of Metabolic Disorders Using NMR-Based Metabolomics: The Shanghai Changfeng Study. Phenomics 1, 186–198 (2021).

6 Surendran, P. et al. Rare and common genetic determinants of metabolic individuality and their effects on human health. Nat Med 28, 2321–2332 (2022).

7 Bartel, J. et al. The Human Blood Metabolome-Transcriptome Interface. PLoS Genet 11, e1005274 (2015).

8 Yin, X. et al. Genome-wide association studies of metabolites in Finnish men identify disease-relevant loci. Nat Commun 13, 1644 (2022).

9 Yin, X. et al. Integrating transcriptomics, metabolomics, and GWAS helps reveal molecular mechanisms for metabolite levels and disease risk. Am J Hum Genet 109, 1727–1741 (2022).

10 Richmond, R. C. & Davey Smith, G. Mendelian Randomization: Concepts and Scope. Cold Spring Harb Perspect Med 12 (2022).

11 Zhao, Q., Wang, J., Hemani, G., Bowden, J. & Small, D. S. Statistical inference in two-sample summary-data Mendelian randomization using robust adjusted profile score. The Annals of Statistics 48, 1742–1769, 1728 (2020).

12 Burgess, S. & Thompson, S. G. Multivariable Mendelian randomization: the use of pleiotropic genetic variants to estimate causal effects. Am J Epidemiol 181, 251–260 (2015).

13 Wang, J. et al. Causal inference for heritable phenotypic risk factors using heterogeneous genetic instruments. PLoS Genet 17, e1009575 (2021).

14 Sun, Y., Lu, Y. K., Gao, H. Y. & Yan, Y. X. Effect of Metabolite Levels on Type 2 Diabetes Mellitus and Glycemic Traits: A Mendelian Randomization Study. J Clin Endocrinol Metab 106, 3439–3447 (2021).

15 Qian, L. et al. Genetically Determined Levels of Serum Metabolites and Risk of Neuroticism: A Mendelian Randomization Study. Int J Neuropsychopharmacol 24, 32–39 (2021).

16 Lord, J. et al. Mendelian randomization identifies blood metabolites previously linked to midlife cognition as causal candidates in Alzheimer’s disease. Proc Natl Acad Sci U S A 118 (2021).

17 Qin, Y. et al. Genome-wide association and Mendelian randomization analysis prioritizes bioactive metabolites with putative causal effects on common diseases. medRxiv (2020).

18 Chen, Y. et al. Genomic atlas of the plasma metabolome prioritizes metabolites implicated in human diseases. Nat Genet 55, 44–53 (2023).

19 Kurki, M. I. et al. FinnGen provides genetic insights from a well-phenotyped isolated population. Nature 613, 508–518 (2023).

20 Davies, N. M., Holmes, M. V. & Davey Smith, G. Reading Mendelian randomisation studies: a guide, glossary, and checklist for clinicians. Bmj 362, k601 (2018).

21 Shu, H. et al. Emerging Roles of Ceramide in Cardiovascular Diseases. Aging Dis 13, 232–245 (2022).

22 Lotta, L. A. et al. Genetic Predisposition to an Impaired Metabolism of the Branched-Chain Amino Acids and Risk of Type 2 Diabetes: A Mendelian Randomisation Analysis. PLoS Med 13, e1002179 (2016).

23 Ferkingstad, E. et al. Large-scale integration of the plasma proteome with genetics and disease. Nat Genet 53, 1712–1721 (2021).

24 Ngo, D., et al. Proteomic profiling reveals biomarkers and pathways in type 2 diabetes risk. JCI Insight 6 (2021).

25 Huang, S. Y. et al. Investigating Causal Relations Between Circulating Metabolites and Alzheimer’s Disease: A Mendelian Randomization Study. J Alzheimers Dis 87, 463–477 (2022).

26 Zhuang, Z. et al. Causal relationships between gut metabolites and Alzheimer’s disease: a bidirectional Mendelian randomization study. Neurobiol Aging 100, 119.e115–119.e118 (2021).

27 Chen, H. et al. Assessing Causal Relationship Between Human Blood Metabolites and Five Neurodegenerative Diseases With GWAS Summary Statistics. Front Neurosci 15, 680104 (2021).

28 Knuplez, E. & Marsche, G. An Updated Review of Pro- and Anti-Inflammatory Properties of Plasma Lysophosphatidylcholines in the Vascular System. Int J Mol Sci 21 (2020).

29 Kinney, J. W. et al. Inflammation as a central mechanism in Alzheimer’s disease. Alzheimers Dement (N Y) 4, 575–590 (2018).

30 Semba, R. D. Perspective: The Potential Role of Circulating Lysophosphatidylcholine in Neuroprotection against Alzheimer Disease. Adv Nutr 11, 760–772 (2020).

31 Hammouda, S. et al. Genetic variants in FADS1 and ELOVL2 increase level of arachidonic acid and the risk of Alzheimer’s disease in the Tunisian population. Prostaglandins Leukot Essent Fatty Acids 160, 102159 (2020).

32 Kovesdy, C. P. Epidemiology of chronic kidney disease: an update 2022. Kidney Int Suppl (2011) 12, 7–11 (2022).

33 Niwa, T. Role of indoxyl sulfate in the progression of chronic kidney disease and cardiovascular disease: experimental and clinical effects of oral sorbent AST-120. Ther Apher Dial 15, 120–124 (2011).

34 Leite, M. I. et al. Myasthenia gravis and neuromyelitis optica spectrum disorder: a multicenter study of 16 patients. Neurology 78, 1601–1607 (2012).

35 Li, X., Sundquist, J. & Sundquist, K. Subsequent risks of Parkinson disease in patients with autoimmune and related disorders: a nationwide epidemiological study from Sweden. Neurodegener Dis 10, 277–284 (2012).

36 Li, Y. & Schellhorn, H. E. New developments and novel therapeutic perspectives for vitamin C. J Nutr 137, 2171–2184 (2007).

37 Gatto, G. J., Jr., Boyne, M. T., 2nd, Kelleher, N. L. & Walsh, C. T. Biosynthesis of pipecolic acid by RapL, a lysine cyclodeaminase encoded in the rapamycin gene cluster. J Am Chem Soc 128, 3838–3847 (2006).

38 Alonso, A. et al. Serum Metabolomics and Incidence of Atrial Fibrillation (from the Atherosclerosis Risk in Communities Study). Am J Cardiol 123, 1955–1961 (2019).

39 Laakso, M. et al. The Metabolic Syndrome in Men study: a resource for studies of metabolic and cardiovascular diseases. J Lipid Res 58, 481–493 (2017).

40 Lim, E. T. et al. Distribution and medical impact of loss-of-function variants in the Finnish founder population. PLoS Genet 10, e1004494 (2014).

41 Zhou, W. et al. Efficiently controlling for case-control imbalance and sample relatedness in large-scale genetic association studies. Nat Genet 50, 1335–1341 (2018).

42 Purcell, S. et al. PLINK: a tool set for whole-genome association and population-based linkage analyses. Am J Hum Genet 81, 559–575 (2007).

43 Mbatchou, J. et al. Computationally efficient whole-genome regression for quantitative and binary traits. Nat Genet 53, 1097–1103 (2021).

