## Supplementary Figures for "Metabolome-wide Mendelian randomization characterizes heterogeneous and shared causal effects of metabolites on human health"

### Supplementary Notes

**Alternative univariable Mendelian randomization**. For each of the 282 metabolite-disease trait pairs that we detected in MR-RAPS at FDR<1%, we evaluated its robustness through testing the causal effect of the metabolite on the disease trait in four alternative MR methods: inverse-variance weighted (IVW) method that assumes no pleiotropy[1], weighted median that assumes ≥50% of the IVs are valid[2], Egger regression that assumes directional pleiotropic effects[3], and Mendelian Randomization Pleiotropy RESidual Sum and Outlier (MR-PRESSO) that identifies horizontal pleiotropic outliers and subsequently excludes outliers from causal estimation[4]. All the analyses were conducted in the TwoSampleMR R package.

**Sensitivity test in Mendelian randomization**. To detect possible violations of MR assumptions, we conducted sensitivity analyses: heterogeneity test between IVs, MR-Egger intercept, MR-PRESSO global, and Steiger filtering tests[5]. Firstly, for MR analyses using ≥ 2 IVs, we conducted Cochran's Q heterogeneity tests to quantify the variability of the causal effect obtained for each separate IV. The Cochran's Q test statistic is expected to follow a chi-squared distribution with degrees of freedom as the number of IVs minus one. Secondly, we estimated MR-Egger intercept to quantify the average pleiotropic effect across IVs[3] and then tested whether the MR-Egger intercept is different from zero. Thirdly, we ran MR-PRESSO global test to detect overall horizontal pleiotropy among IVs through comparing the observed distance between all the IVs and the regression line of causal effect to the expected distance under the null hypothesis of no horizontal pleiotropy[4]. Lastly, we applied a Steiger filtering test to infer the causal direction. For each IV, Steiger filtering test evaluated whether it explains more variance for disease traits than for metabolites.

### References


1. Burgess S, Butterworth A, Thompson SG. Mendelian randomization analysis with multiple genetic variants using summarized data. **Genet Epidemiol**. 2013;37(7):658-65.
2. Bowden J, Smith GD, Haycock PC, Burgess S. Consistent Estimation in Mendelian Randomization with Some Invalid Instruments Using a Weighted Median Estimator. **Genet Epidemiol**. 2016;40(4):304-14.
3. Bowden J, Smith GD, Burgess S. Mendelian randomization with invalid instruments: effect estimation and bias detection through Egger regression. **Int J Epidemiol**. 2015;44(2):512-25.
4. Verbanck M, Chia-Yen Chen CY, Neale B, Do R. Detection of widespread horizontal pleiotropy in causal relationships inferred from Mendelian randomization between complex traits and diseases. **Nat Genet**. 2018;50(5):693-698.
5. Hemani G, Tilling K, Smith GD. Orienting the causal relationship between imprecisely measured traits using GWAS summary data. **PLoS Genet**. 2017;13(11):e1007081.

**Supplementary Figure 1: Number of instrumental variables for each metabolite**

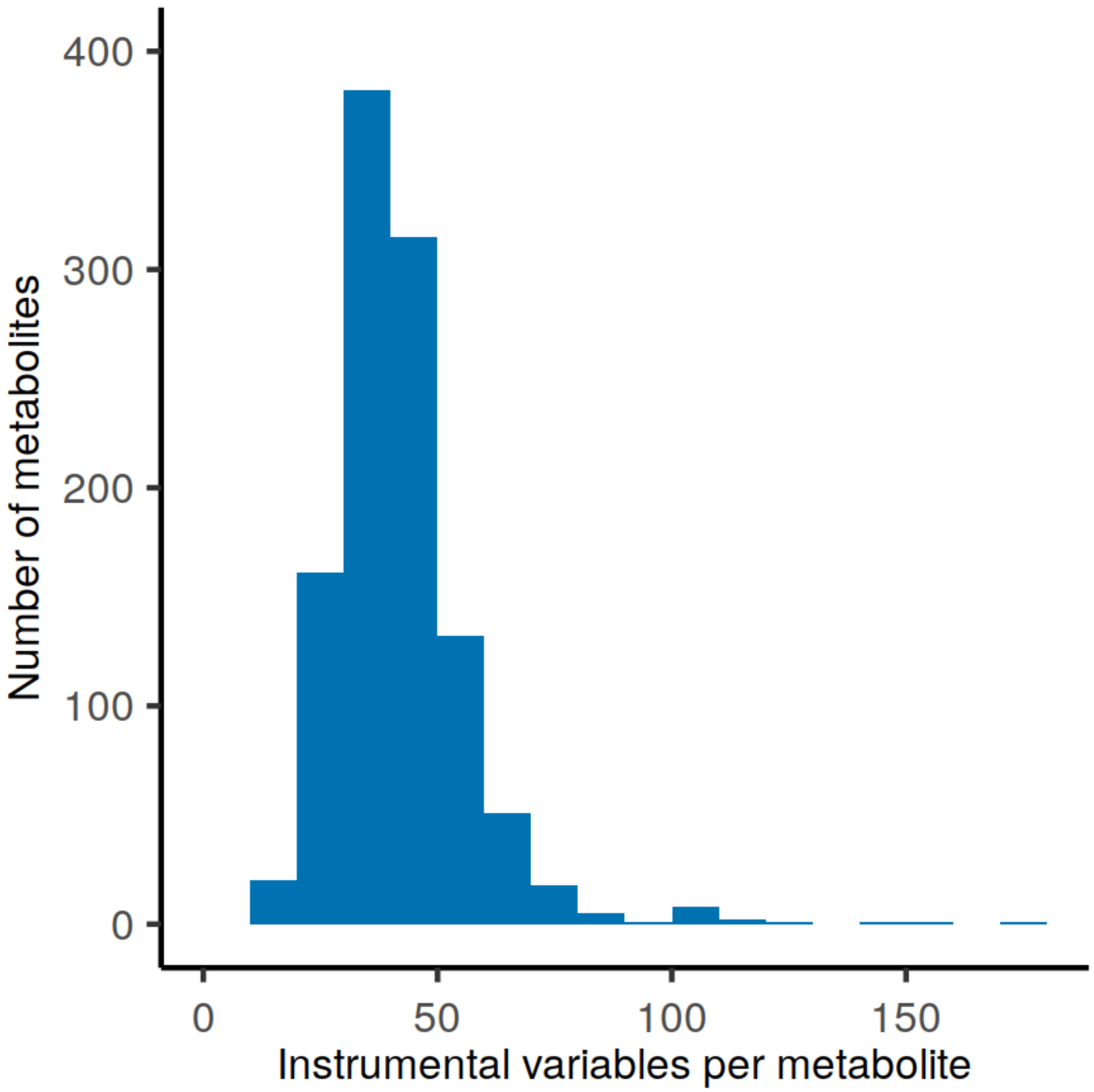

**Supplementary Figure 2: Results of the 282 causal effects identified by MR-RAPS in four alternative Mendelian randomization approaches.** Egger: MR-Egger regression; WM: Weighted median; IVW: inverse variance weighted Mendelian randomization; MR_PRESSO: Mendelian Randomization Pleiotropy RESidual Sum and Outlier

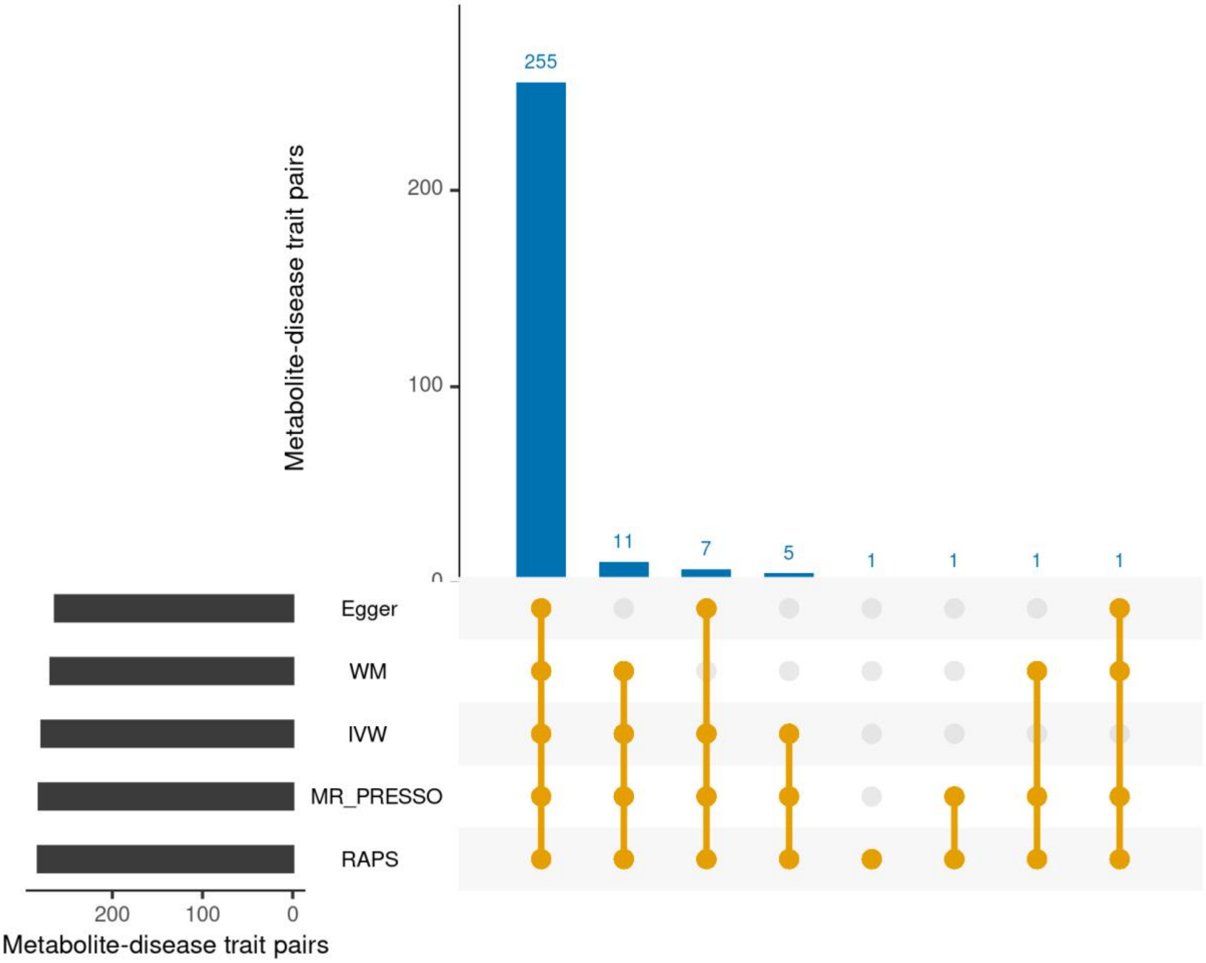

**Supplementary Figure 3: Distribution of *p*-values for Q statistics in the heterogeneity test for the 282 causal associations.** The vertical red light denotes the *p*-value threshold of 0.05.

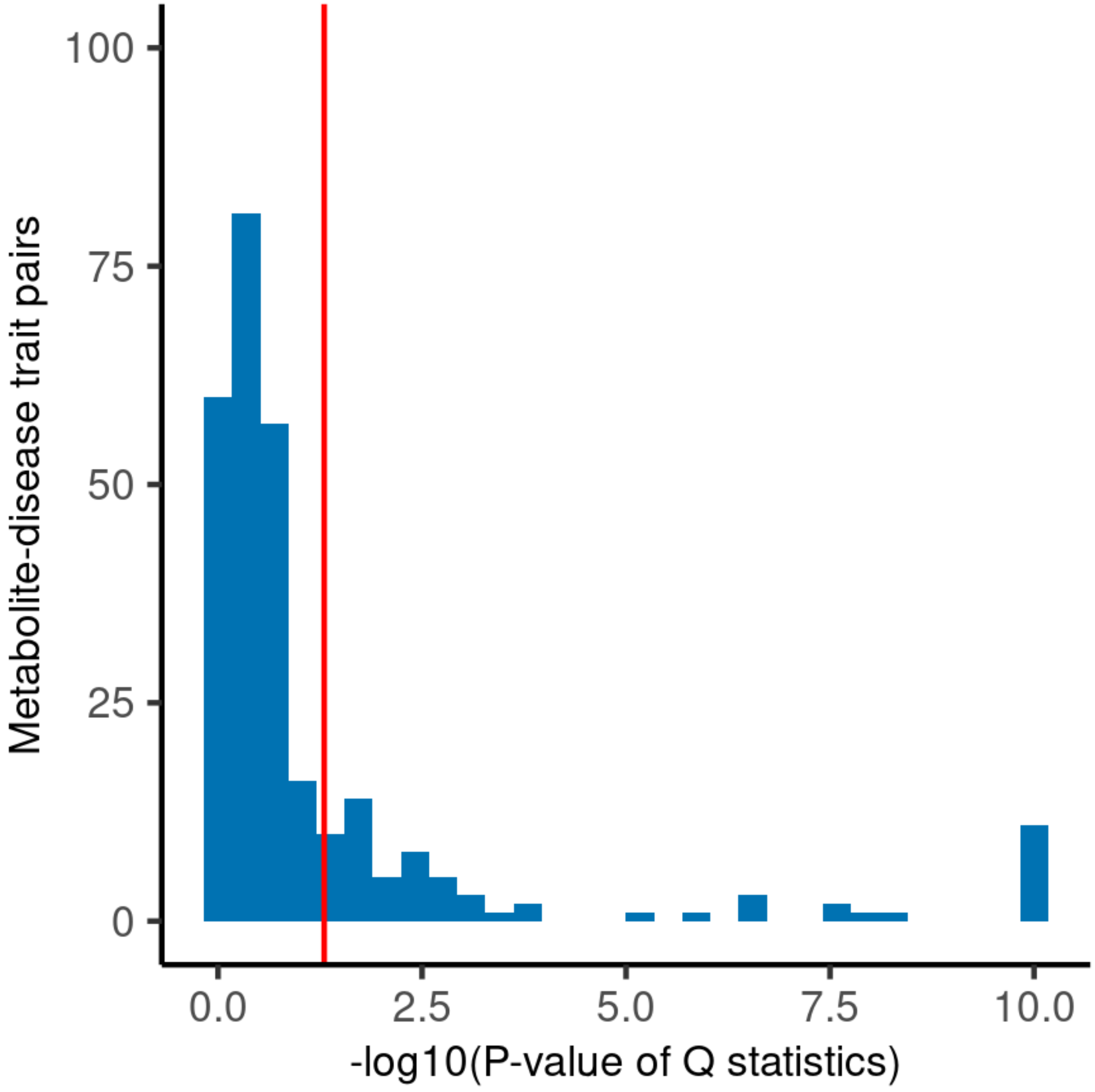

**Supplementary Figure 4: Distribution of *p*-values for Egger intercept for the 282 causal associations.** The vertical red light denotes the *p*-value threshold of 0.05.

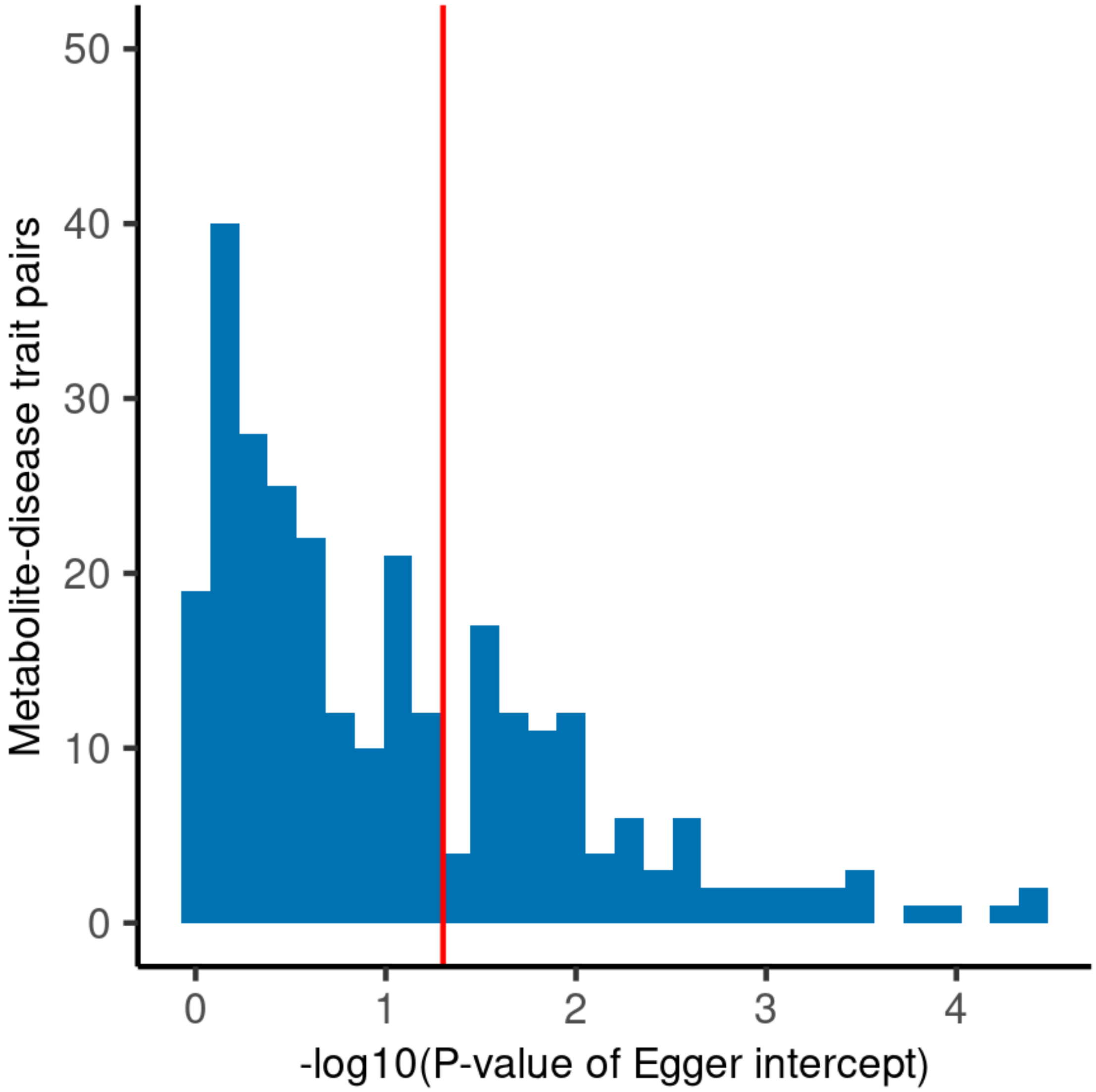

**Supplementary Figure 5: Distribution of *p*-values for MR-PRESSO global test for the 282 causal associations.** The *p*-values are capped at $10^{-10}$. The vertical red light denotes the *p*-value threshold of 0.05.

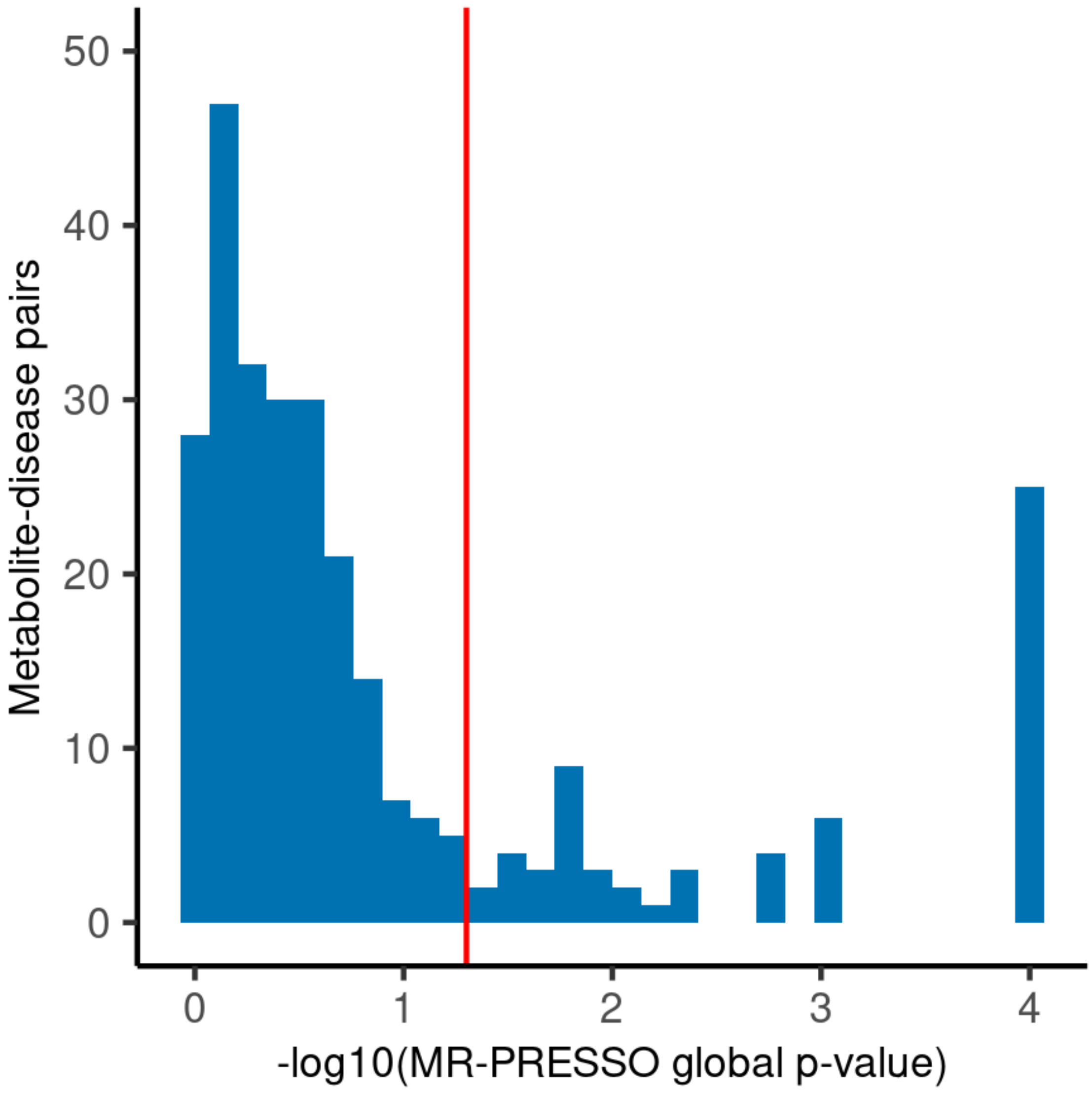

**Supplementary Figure 6: Heatmap of (a) phenotypic Pearson correlation and (b) correlation of instrumental variables (IV correlation) between the three N-acyl-alpha amino acids**

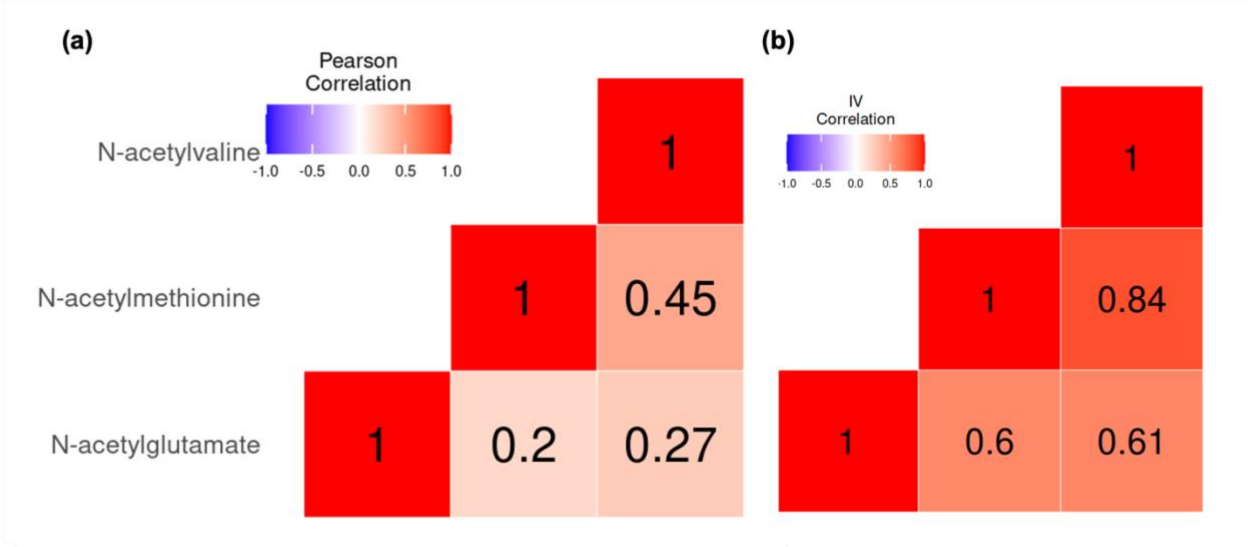

### Supplementary Figure 7: Heatmap of phenotypic Pearson correlations between all pairs of the 70 metabolites

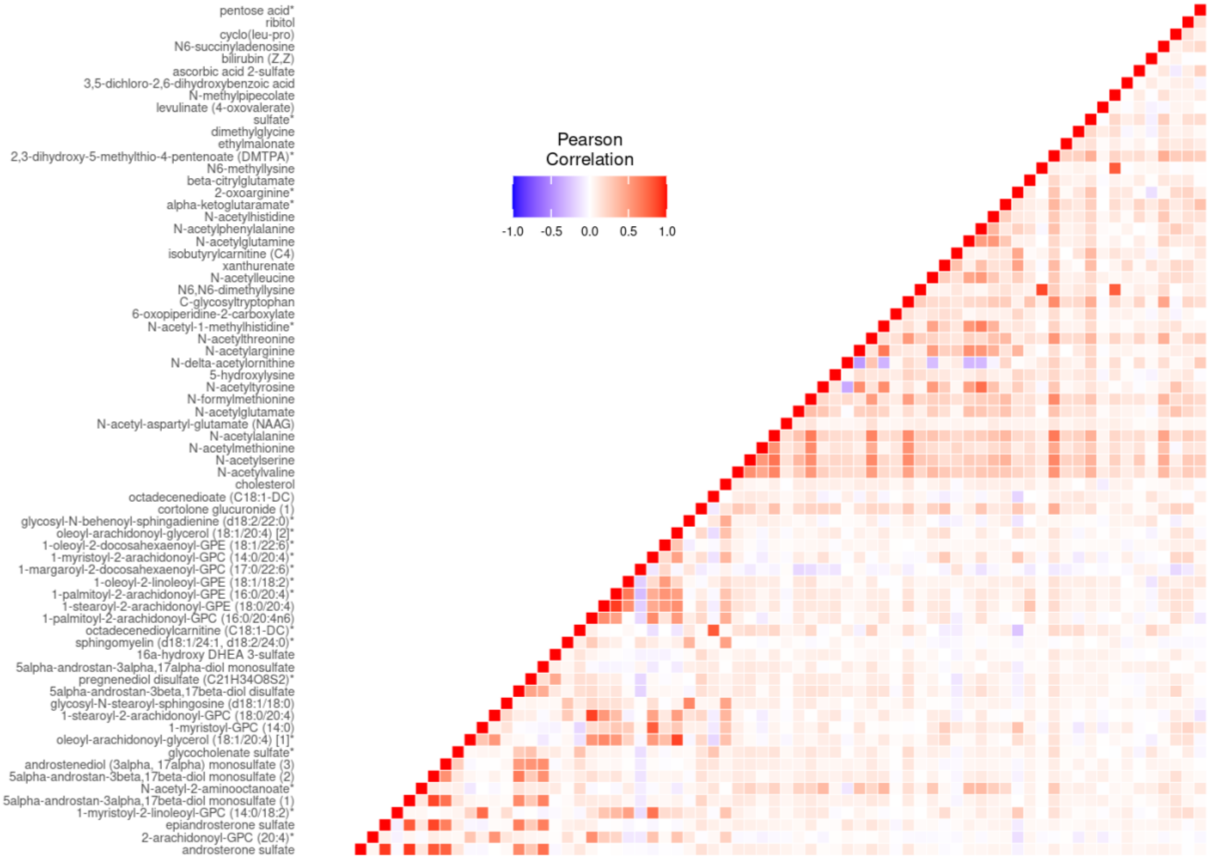

**Supplementary Figure 8: Comparison of phenotypic and instrumental variable (IV) correlations between all pairs of the 70 metabolites**

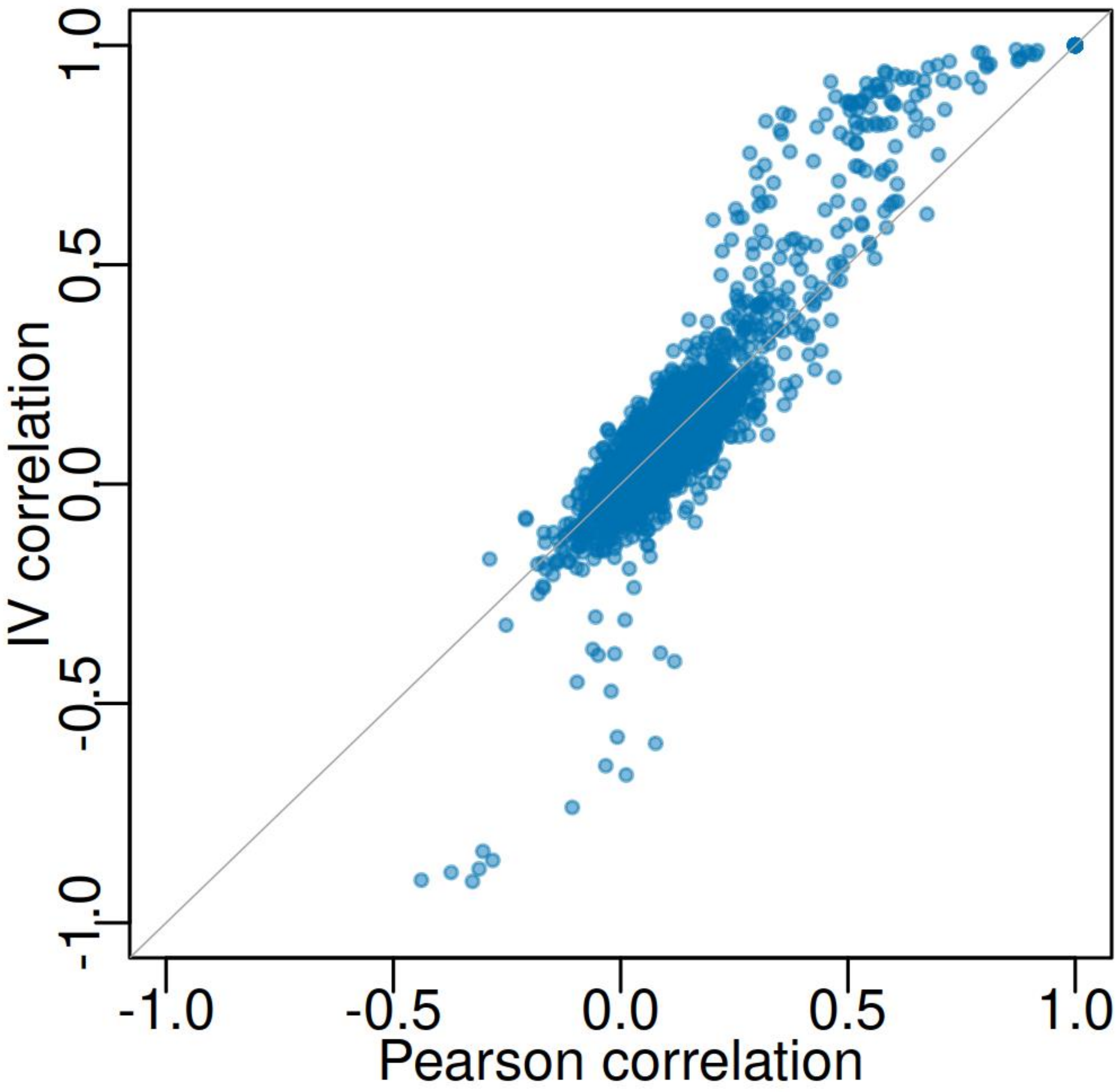

**Supplementary Figure 9: Distribution of pairwise instrumental variable (IV) correlations between metabolites with significant causal effects on the same disease trait**

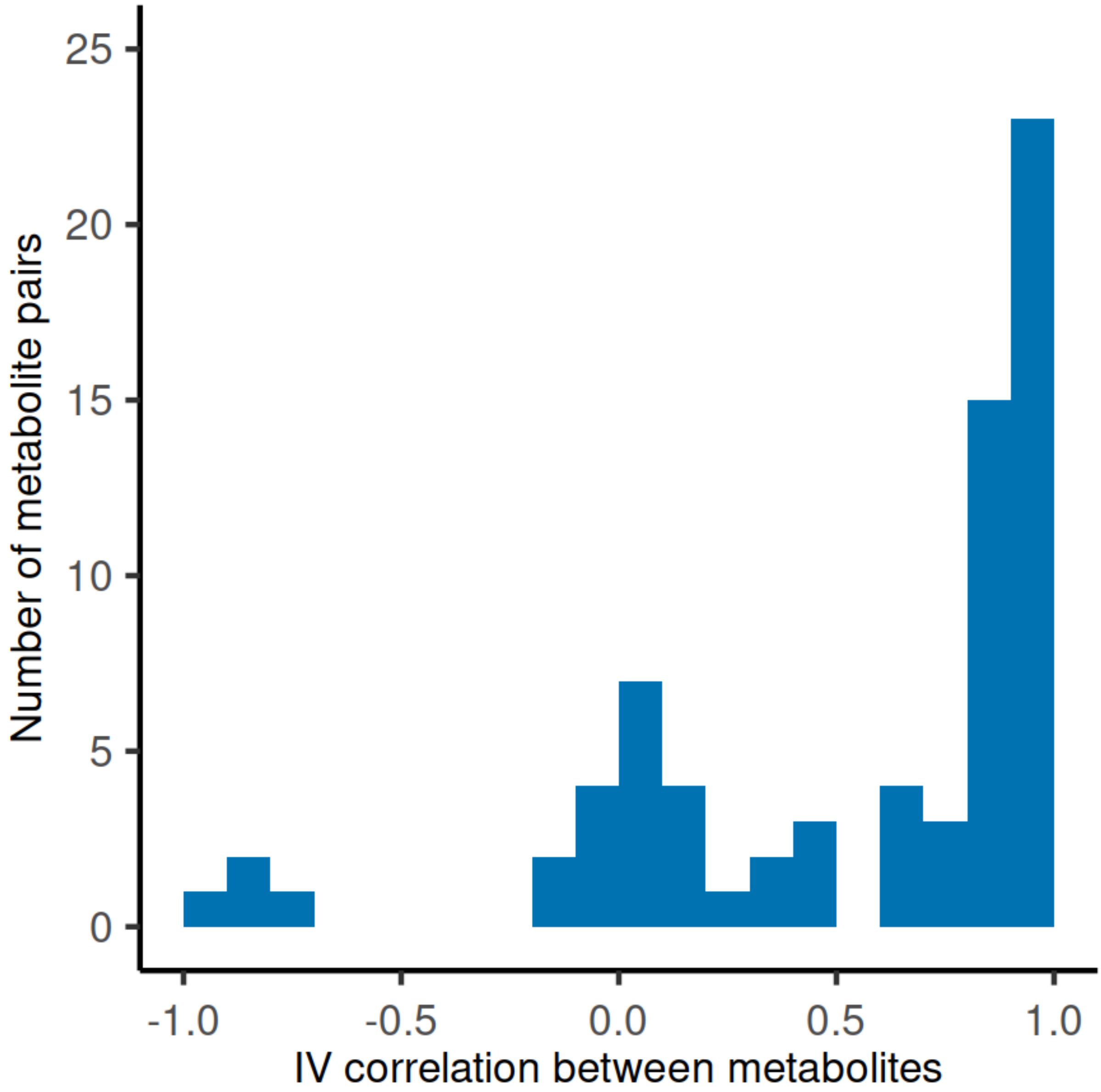

**Supplementary Figure 10: Genetic associations for the ratio of N6,N6-dimethyllysine and N-methylpipecolate at (a) *NAT8*; (b) *SLC6A20*; (c) *AKR1C1/2/3/4/8*; (d) *PYROXD2*; and (f) *SLC7A9*.**

**(a) the *NAT8* region**

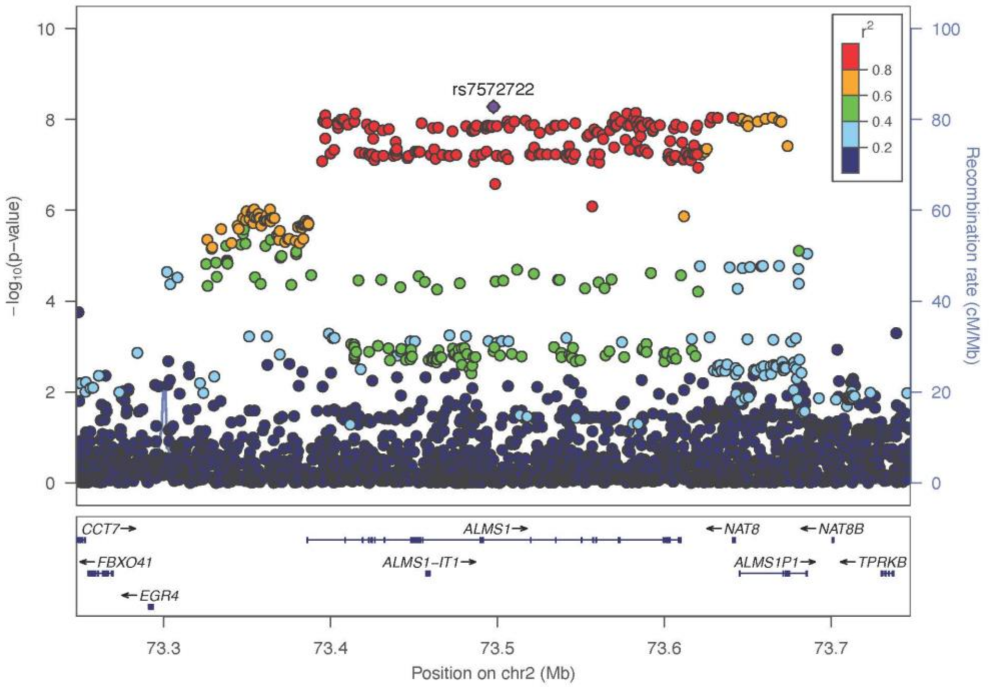


**(b) the *SLC6A20* region**

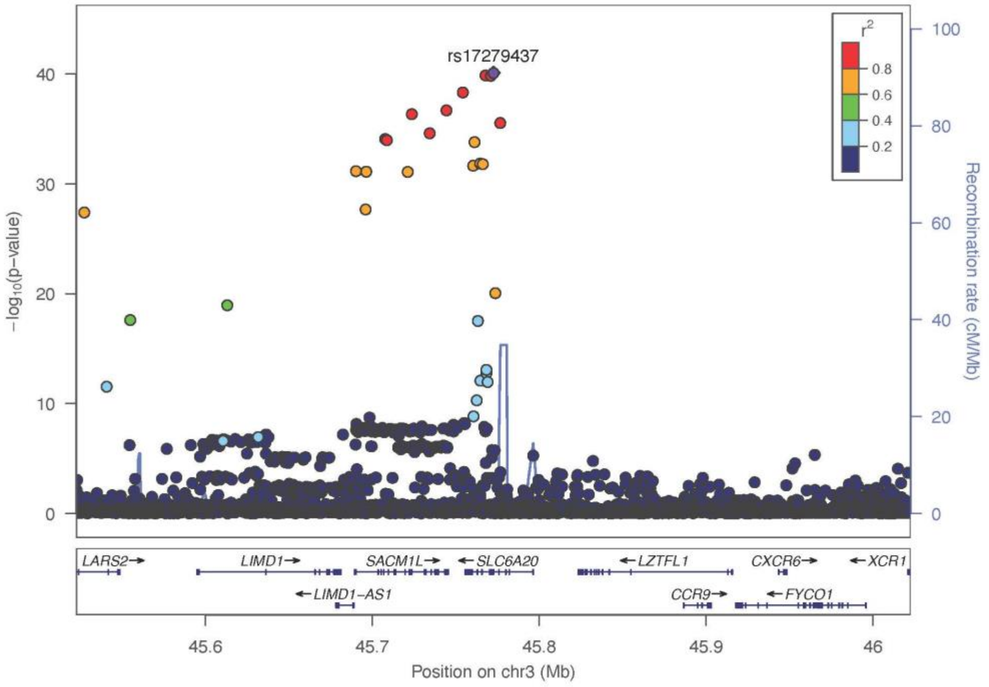

**(c) the *AKR1C1/2/3/4/8* region**

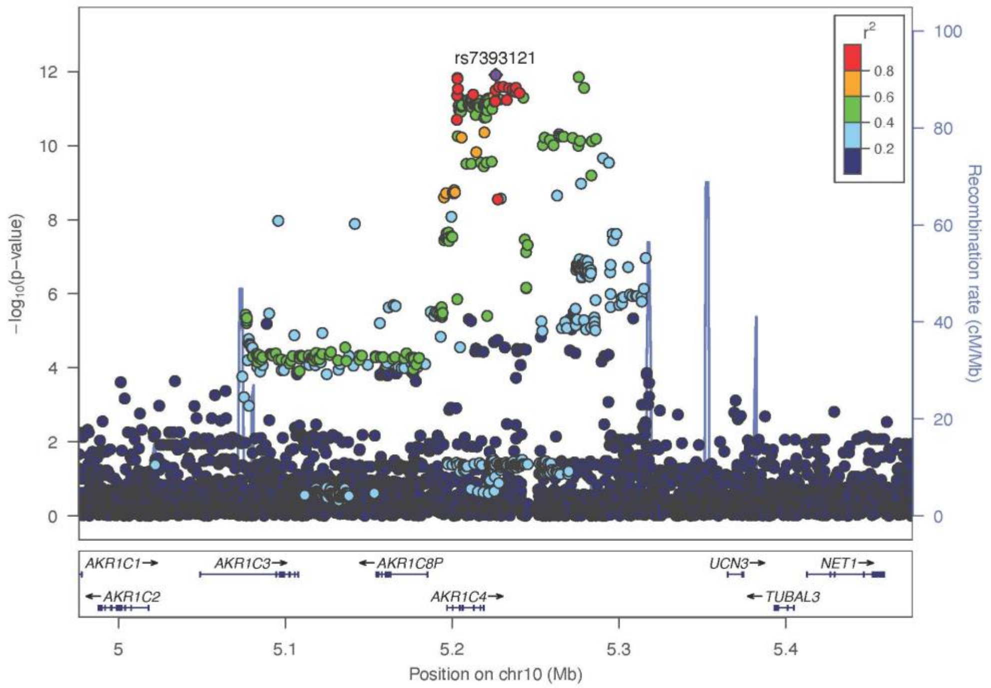


**(d) the *PYROXD2* region (two independent signals)**

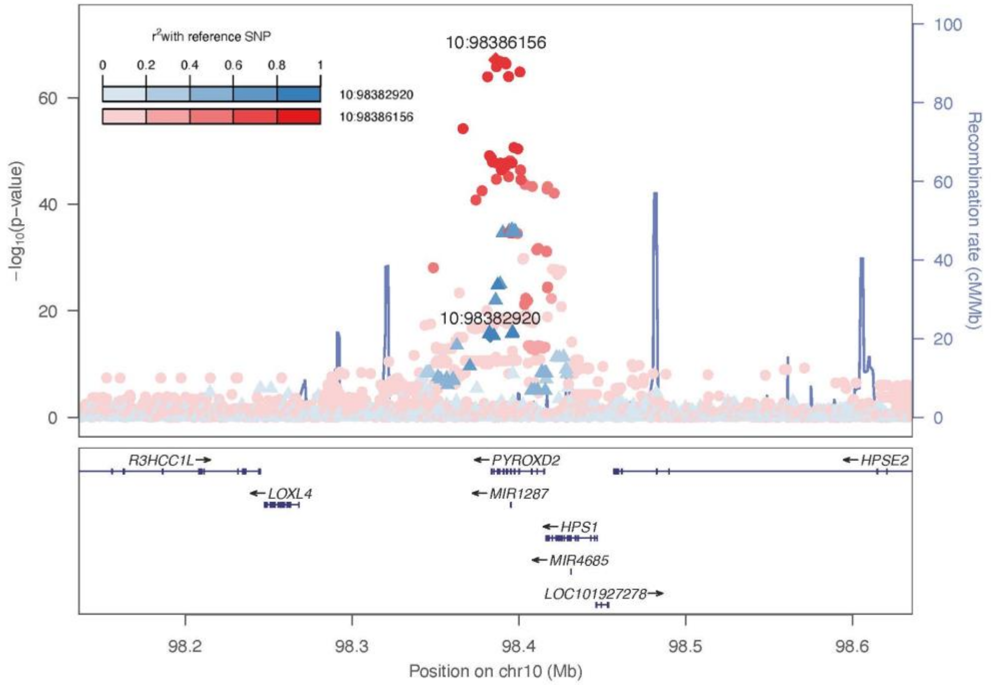

**(e) the *SLC7A9* region (two independent signals)**

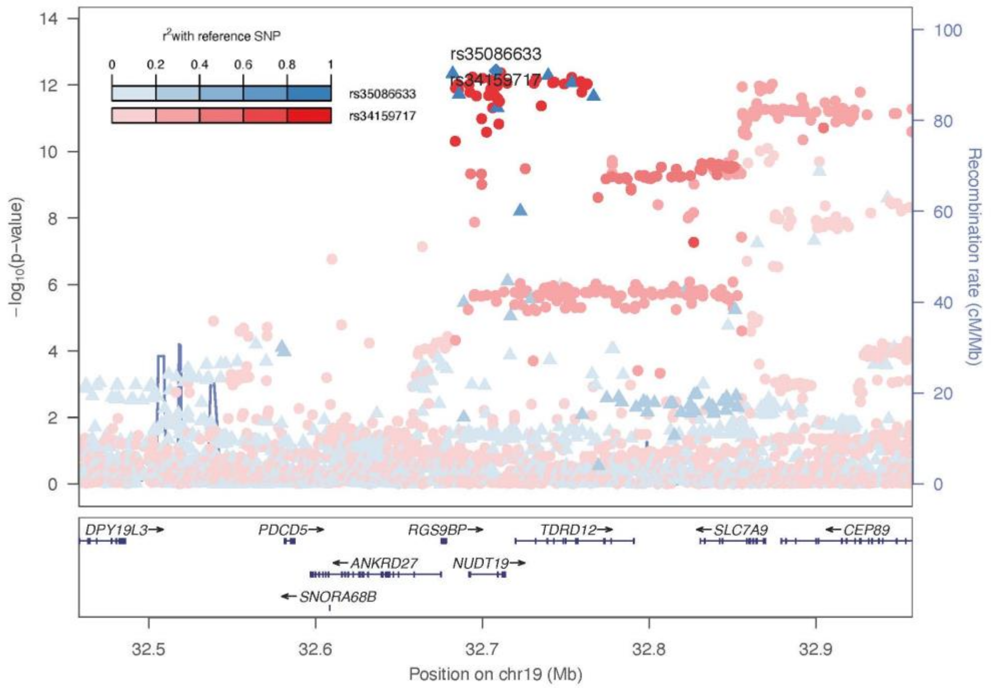

**Supplementary Figure 11: Causal effect of metabolite ratio between N6,N6-dimethyllysine and N-methylpipecolate on risk of anxious personality disorder : (a) scatter plot of effects of the five instrumental variables on metabolite ratio (X-axis) and anxious personality disorder (Y-axis). The slope of the blue line denotes the causal effect estimated in MR-RAPS; (b) forest plot of causal effects on anxious personality disorder.** Each dot denotes one independently-associated variant.

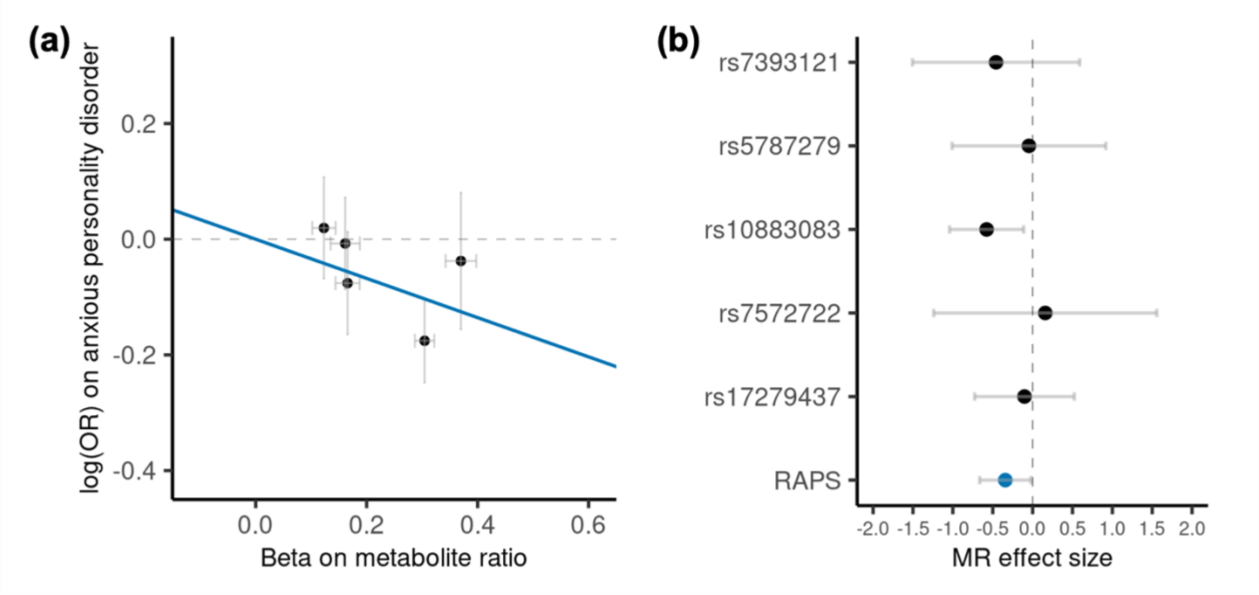
